# Brain activity as a candidate biomarker for personalised caffeine treatment in premature neonates

**DOI:** 10.1101/2025.05.02.25326856

**Authors:** Fatima Usman, Coen S. Zandvoort, Shellie Robinson, Mariska Peck, Maria M. Cobo, Tricia Adjei, Luke Baxter, Ria Evans Fry, Annalisa G. V. Hauck, Richard Rogers, Gabriela Schmidt Mellado, Alexandra Scrivens, Marianne van der Vaart, Maarten De Vos, Eleri Adams, John van den Anker, Caroline Hartley

**Affiliations:** Department of Paediatrics, University of Oxford, Oxford, UK; Universidad San Francisco de Quito USFQ, Colegio de Ciencias Biologicas y Ambientales, Quito, Ecuador; Nuffield Division of Anaesthetics, John Radcliffe Hospital, Oxford University Hospitals NHS Foundation Trust, Oxford, UK; Department of Electrical Engineering (ESAT), STADIUS Center for Dynamical Systems, Signal Processing and Data Analytics, KU Leuven, Leuven, Belgium; Department of Development and Regeneration, University Hospitals Leuven, Child Neurology, KU Leuven, Leuven, Belgium; Newborn Care Unit, John Radcliffe Hospital, Oxford University Hospitals NHS Foundation Trust, Oxford, UK; Division of Clinical Pharmacology, Children’s National Hospital, Washington DC, USA

## Abstract

**Background:** Medication in hospitalised infants is often prescribed using a ‘one-size-fits-all’ approach due to lack of clinical biomarkers. Caffeine is one of the most frequently administered medicines in neonatology - prescribed for the management of apnoea of prematurity, to aid extubation and increasingly for conditions such as bronchopulmonary dysplasia. Caffeine guidelines for the management of apnoea of prematurity indicate use based on the age of the infant, but this does not account for individual variation in apnoea rate. Consequently, infants may risk caffeine undertreatment or adverse events due to over-exposure. Apnoea in preterm infants is related to nervous system immaturity, hence, as an essential first step to assess whether brain activity may be a useful biomarker for caffeine treatment, we tested the hypothesis that apnoea rate is related to brain activity.

**Methods and Findings:** In this single-centre prospective observational cohort study, we simultaneously recorded brain activity using electroencephalography (EEG) and respiration using impedance pneumography in 74 infants aged 31-36 weeks postmenstrual age (PMA) on 138 separate occasions. We demonstrate that apnoea rate in moderate/late preterm infants is dependent on brain age gap (defined as the difference between the infant’s age assessed from their brain activity and their PMA). In contrast, apnoea rate is not correlated with PMA in this age range. In an exploratory sub-study, we provide initial evidence that when caffeine is discontinued, infants with immature brain activity have more frequent apnoeas and desaturations compared with those with more mature brain function.

**Conclusions:** These findings provide initial evidence to indicate that brain age gap (assessed automatically using machine learning) is a candidate biomarker for personalised caffeine treatment in preterm infants.

## Introduction

Apnoea of prematurity (AOP) is a developmental respiratory disorder affecting more than half of the 15 million infants born prematurely each year worldwide (1,2). Apnoea is a breathing cessation of variable duration which can be life-threatening and may impact long-term neurodevelopmental outcomes (3,4), so it is essential to optimise treatment. AOP is a consequence of nervous system and pulmonary immaturity of preterm infants (5) and will usually resolve by late prematurity (6).

Caffeine is the primary pharmacological treatment for AOP, reducing apnoea rates, duration of ventilatory support and extubation failure, and improving neurodevelopmental outcomes (7). Clinical guidelines for the treatment and prevention of AOP indicate caffeine requirement based on gestational age or weight of an infant. For example, the World Health Organisation recommends that all infants born before 34 weeks’ gestation are given caffeine (8). Similarly, the European Consensus Guidelines suggests to ‘use caffeine routinely in infants <32 weeks of gestation’ (9) and the UK’s National Institute for Health and Care Excellence (NICE) guidelines state ‘use caffeine citrate routinely in preterm babies born at or before 30 weeks, starting it as early as possible and ideally before 3 days of age’ (10). Moreover, NICE guidelines also suggest to ‘consider stopping caffeine citrate at 33 to 35 weeks’ corrected gestational age if the baby is clinically stable.’ Whilst this does take into account the stability of the baby, it nevertheless also relates to age, precluding stopping treatment before 33 weeks. Prescription based on age does not directly relate to the underlying cause of apnoea (i.e., immaturity of the brain and pulmonary systems) (Figure 1). This may expose some infants to caffeine-related adverse effects, such as tachycardia, reflux and feeding intolerance, or rarely seizures (11,12) if, for example, they are born just before the cut-off age and receive caffeine unnecessarily. Conversely, some infants born just after the cut-off age, who are relatively neurologically immature, and who may benefit from treatment, may not receive it immediately. Moreover, some infants discontinue treatment too early and experience substantial apnoeas necessitating recommencement of treatment (13). An individualised approach is needed (14) and can be achieved through identification of biomarkers for caffeine requirement. Currently, no such biomarkers have been identified.

**Figure 1.**
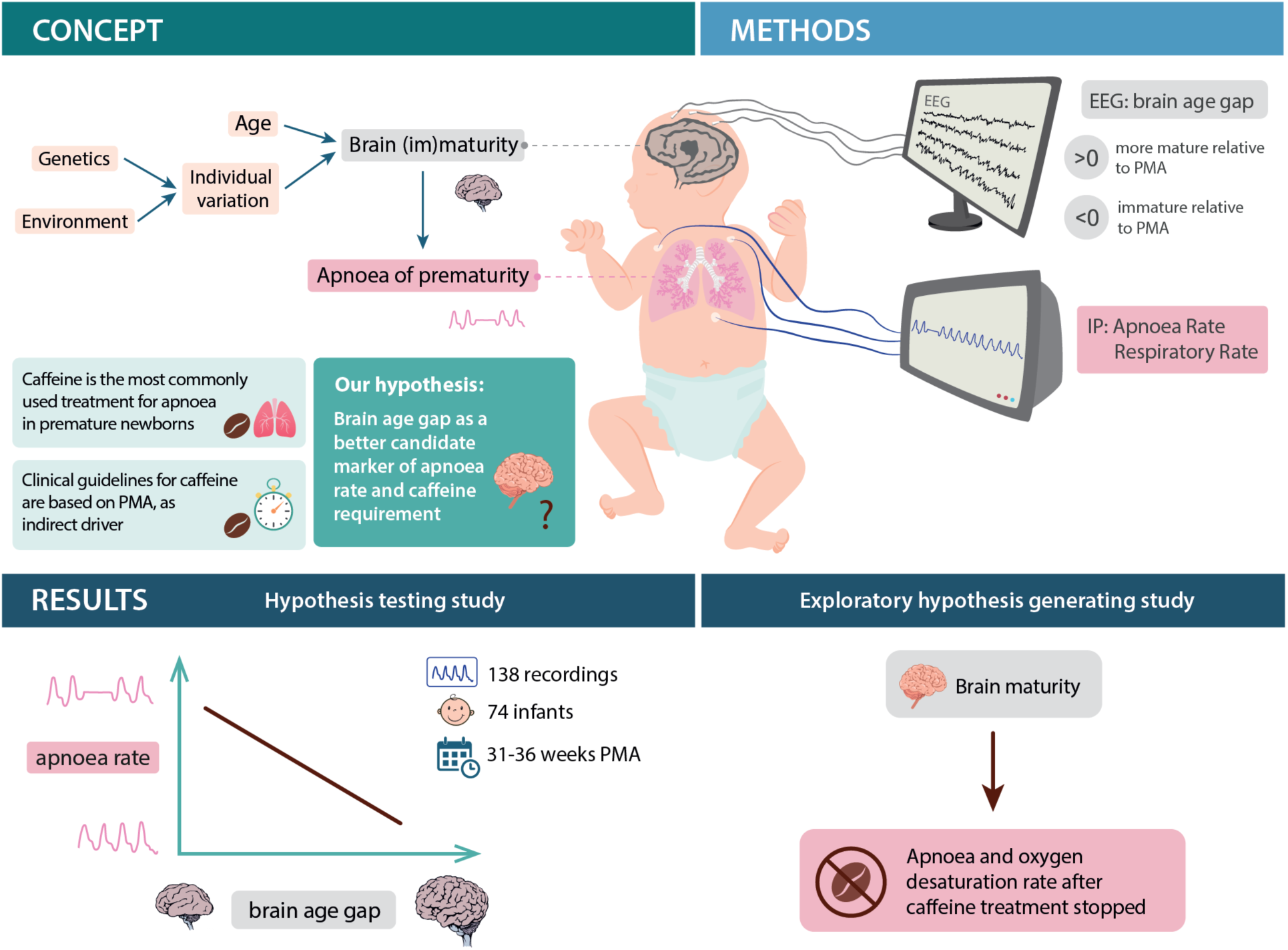
Schematic of study concept, design and main results. EEG: electroencephalography, IP: impedance pneumography, PMA: postmenstrual age. Note that other factors, such as infection, can lead to apnoea in premature infants; however, here we focus on the relationship between apnoea and brain development.

Given the direct link between brain development and apnoea, here we propose brain activity, and more specifically, a metric derived from brain activity known as the brain age gap, as a candidate biomarker (Figure 1). We define brain age gap as the difference between an infant’s ‘brain age’ and their postmenstrual age (PMA) (15,16). Thus, the brain age gap is a marker of whether an individual’s brain activity is more or less mature relative to their PMA, with negative values of brain age gap indicating that their brain activity is immature compared with their PMA and positive values indicating that their brain activity is more mature than their PMA. Brain age can be assessed using electroencephalography (EEG), combined with recently developed and validated machine learning models, and uses features in the EEG compared with the model training data to ascertain an infant’s brain age (17–20). These models capitalise on the fact that the EEG changes rapidly in preterm infants – the EEG of a very preterm infant is characterised by discontinuous activity interspersed with high amplitude bursts. Brain activity becomes increasingly continuous and lower in amplitude with increasing age (21). Similarly, sensory-evoked responses change with PMA (22–24). Very immature brain activity relative to PMA may be associated with poorer later life neurodevelopmental outcomes (17,18), yet there will also be normal variation in brain age gap – just as older children learn to walk and talk at different ages, brain developmental trajectories exhibit individual variation modulated by factors such as genetics and the environment (25,26). We postulate that this individual variation in brain age gap (i.e., individual variation in EEG activity) will explain variation in apnoea rate and consequently could be used to guide treatment with caffeine.

There is limited evidence to date to suggest that brain activity in human preterm infants is related to apnoea rate: Henderson-Smart and colleagues (27) demonstrated prolonged auditory brainstem conduction times in infants with AOP. Moreover, we recently demonstrated that cortical brain activity (most likely originating from cortical motor areas) is involved in the regulation of breathing in infants and that infants with stronger ‘coupling’ between cortical and respiratory activity had lower incidences of apnoea (28). In the present study, we hypothesised that, in late preterm infants, apnoea rate is related to brain age gap and that this relationship is stronger than the relationship between apnoea rate and PMA. Indeed, demonstrating that there is a significant correlation between apnoea rate and brain age gap, and that this is stronger than the relationship between apnoea rate and PMA, is an essential first step to suggest that brain age gap may be a useful biomarker for caffeine treatment; if this were not the case then treatment based on an alternative biomarker would be a better approach. For this study, we focused on moderate-to-late preterm infants from 31–36 weeks PMA to investigate whether the brain age gap can be used to assess caffeine requirement. It is possible that some of even the youngest infants in this age range will not need to start caffeine treatment and that brain age gap could be used to guide this decision. Moreover, brain age gap may be a candidate biomarker to indicate when to discontinue treatment in those who have received it, which is an important step in their care, expedites discharge and could shift the balance in an individual from adverse effects and towards benefit.

## Results and Discussion

We included 74 infants on 138 separate occasions (demographic information is provided in Table 1). We included preterm infants between 31 – 36 weeks PMA who were stable and without any significant respiratory or central nervous system complications at the time of the EEG recording (additional details in the Methods). EEG and vital signs were recorded for one to two hours, with continued vital signs recording in some infants up to the time of discharge (see Methods). Sensory-evoked (visual and tactile) and resting state EEG were used to calculate the brain age gap by applying previously developed and validated models (17,18). We used both sensory-evoked and resting state models to improve accuracy by assessing these different (i.e. sensory-evoked and resting state) aspects of functional brain age gap; results using each model independently are presented in the supplementary material and demonstrate similar trends to those presented in the main results (Figure S8).

**Table 1.**
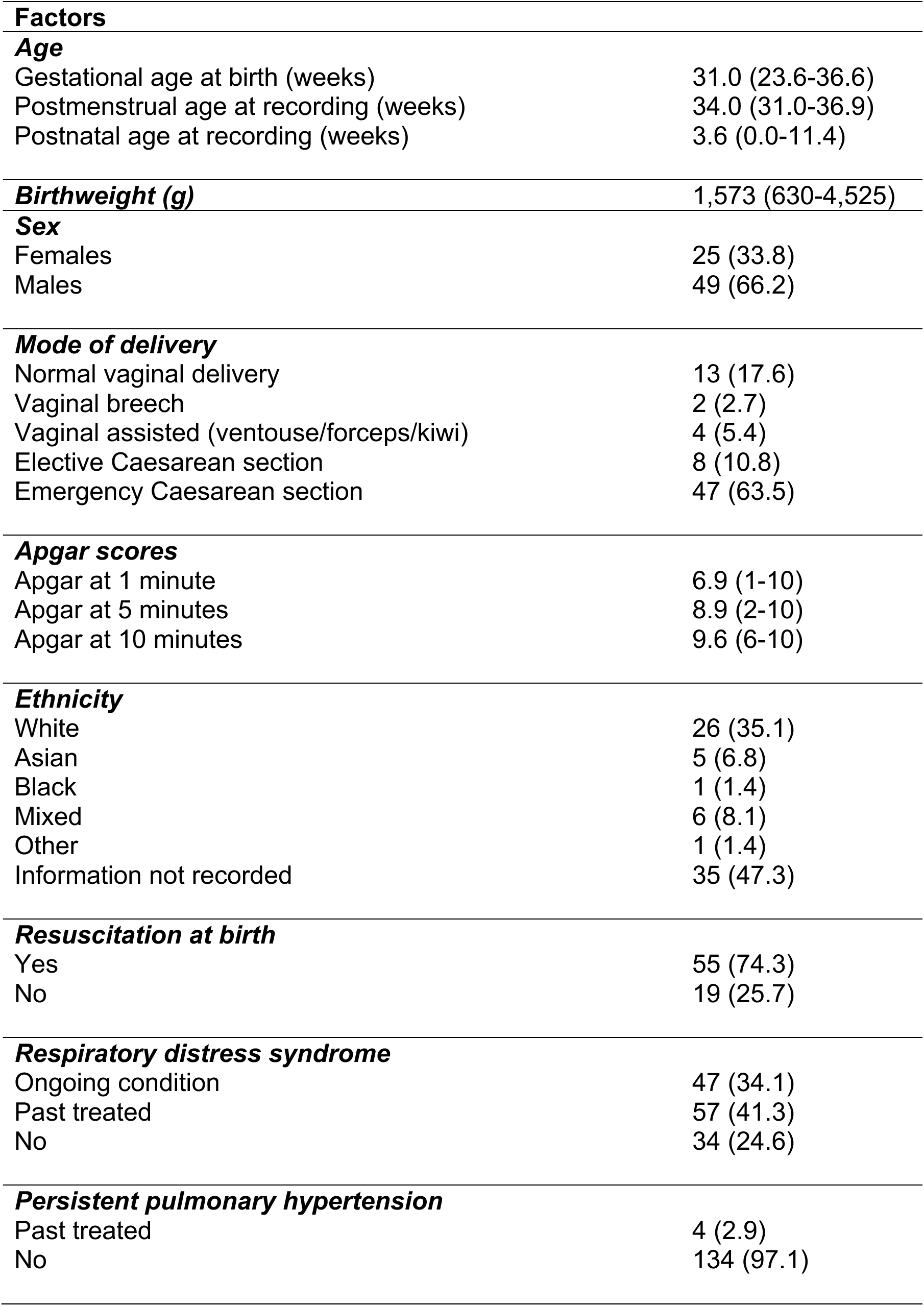
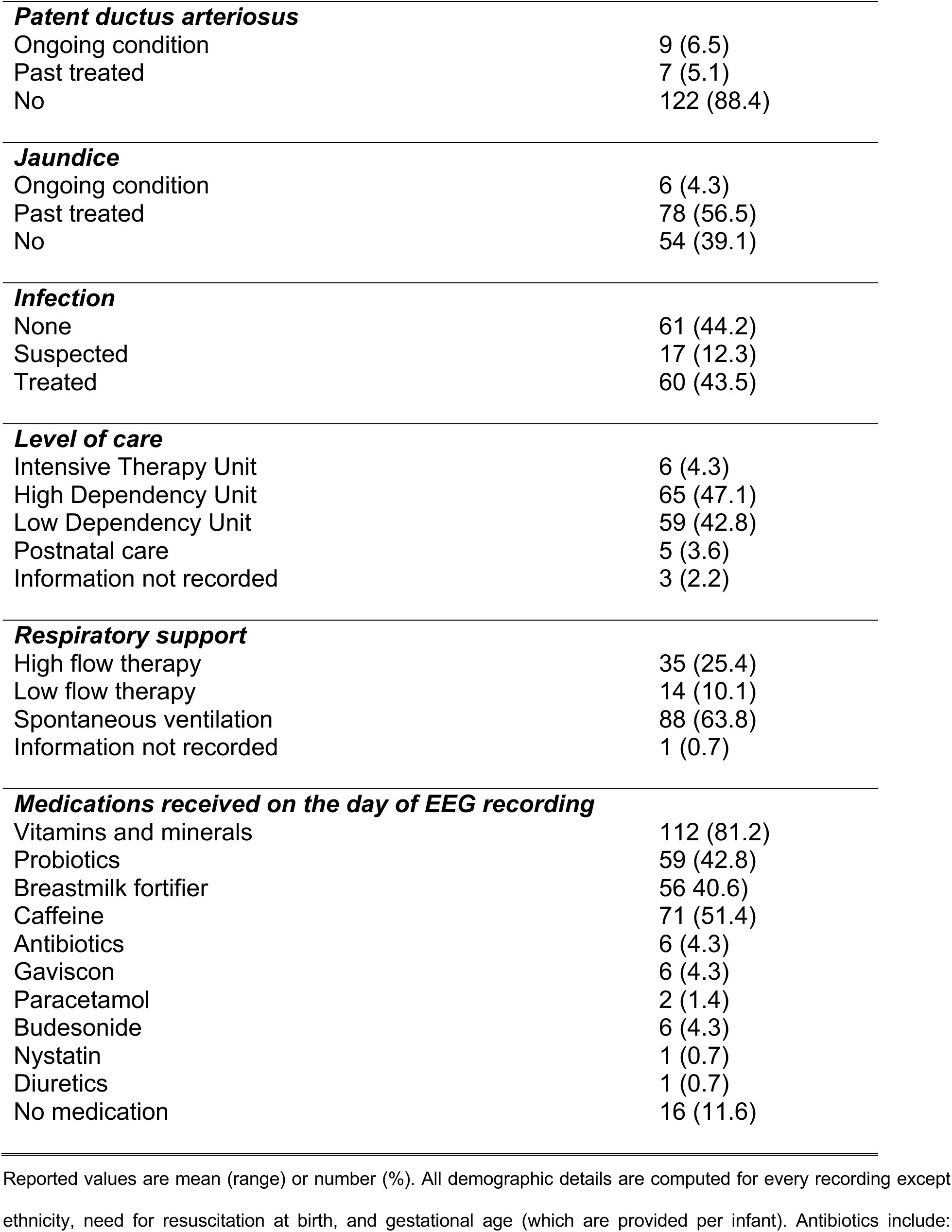
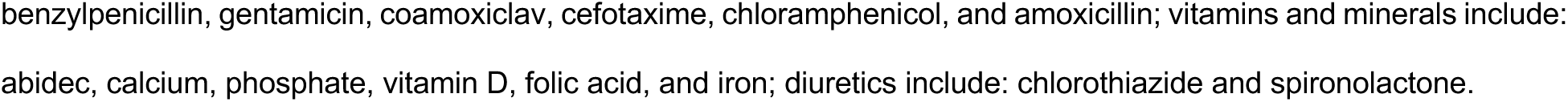
Infant demographics.

Apnoea rate was not significantly associated with PMA in the age range studied (*p*:0.58; β [95% CI]:-0.04 [−0.16; 0.09]; ρ:-0.0472; E_point_:1.26; E_CI_:1.00; Figure 2A). Whilst we would expect that apnoea rate decreases with PMA when considering a wider age range (6), this result demonstrates that apnoea rate is not strongly related to PMA in older preterm infants, highlighting potential pitfalls of guidelines based on age. Consistent with our hypothesis, apnoea rate was significantly related to brain age gap (*p*:0.024; β [95% CI]:- 0.22 [−0.41; −0.03]; ρ:-0.19; E_point_:1.83; E_CI_:1.22; Figure 2B, comparison of the relationship between apnoea rate and brain age gap versus apnoea rate and PMA: *p*:0.12; one-tailed bootstrapped 95% CI: [−0.2287, 1]). There was a clear relationship between brain age gap and the distribution of prolonged inter-breath intervals, with immature brain activity associated with longer inter-breath intervals (Figure 2C), suggesting the association between apnoea rate and brain age gap holds irrespective of the exact definition of apnoea used, which is variable within clinical practice (29). This relationship between apnoea rate and brain age gap was also observed in the subgroup of infants receiving caffeine at the time of study (Supplementary Figure S9), providing preliminary indications of the potential value of brain age gap as a biomarker for caffeine requirement. In preterm infants, the development of central chemoreceptors is immature, and there is a blunted central nervous system response to hypercapnia (30). This immaturity of respiratory centres, which we postulate is related to general immaturity of the brain as assessed here, likely underlies these results. Moreover, these results may be related to immaturity of the cortical control of respiration (28).

**Figure 2.**
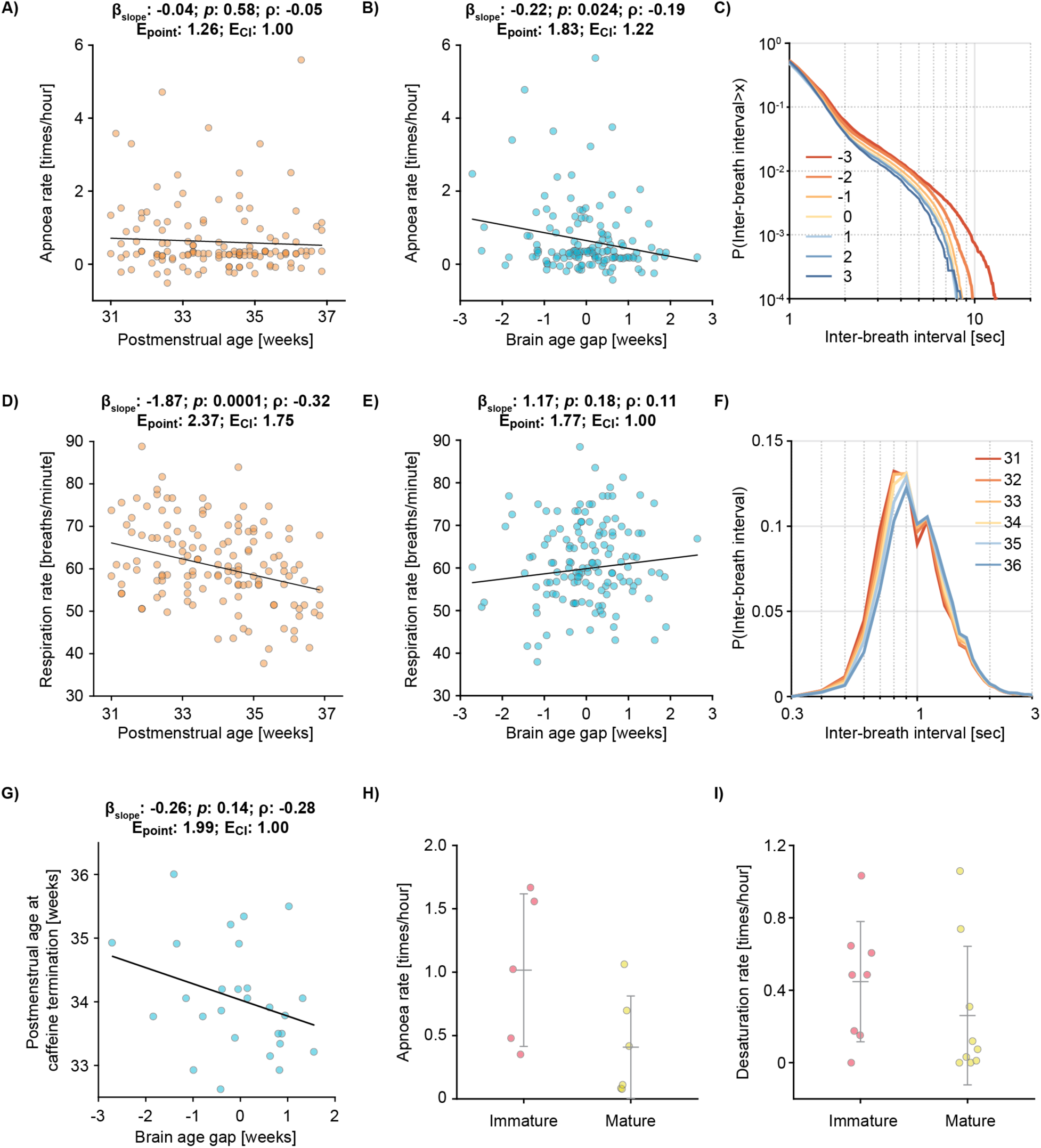
Apnoea rate and caffeine requirement are related to brain age gap. The relationship between apnoea rate (defined as a pause in breathing of at least 15 seconds) and (A) postmenstrual age (PMA), and (B) brain age gap. Brain age gap is computed relative to the infant’s PMA, with negative values indicating that the brain activity is immature relative to PMA. Each dot indicates an individual test occasion (A-F, 74 infants were included on 138 test occasions). Black line is the mean fit of the linear model (β_slope_, *p*, ρ, E_point_, and E_CI_ in the titles are the predictor’s regression slope, and its significance, partial correlation coefficient, E-value point estimate, and E-value of the confidence interval, respectively). (C) The cumulative probability distribution of inter-breath intervals split according to brain age gap. The distribution is shown on a double logarithmic scale to emphasise the prolonged inter-breath intervals (i.e., apnoeas). (D, E) The relationship between respiratory rate and (D) PMA and (E) brain age gap. (F) The probability density distributions of inter-breath intervals shown according to the infant’s PMA. (G) The relationship between age at which caffeine treatment is discontinued (dependent on clinical decision) compared to brain age gap in the subset of 27 infants where brain activity was recorded in the two weeks before caffeine was discontinued. (H, I) Exploratory hypothesis-generating study in the subset of infants where vital signs were recorded longitudinally. Infants were grouped according to whether their brain activity was immature or mature relative to their PMA before they discontinued caffeine treatment. Error bars indicate mean and standard deviation. (H) Apnoea rate (n = 11 infants) and (I) desaturation rate (n = 17 infants) were computed in the 7 days after treatment was stopped. Statistical models in panels A and B are adjusted for data length of impedance pneumograph (i.e. the signal used to assess apnoea rate) and infection; panels D, E and G are adjusted for infection (see Supplementary Results for discussion of statistical models and Supplementary Figures S10-S16).

Interestingly, when considering the average respiratory rate (i.e., normal breathing between apnoeas) we observed the opposite results - respiratory rate significantly decreased with PMA (*p*:0.0001; β [95% CI]:-1.87 [−2.81; −0.93]; ρ:-0.3186; E_point_:2.37; E_CI_: 1.75; Figure 2D, F) and respiratory rate did not significantly relate to brain age gap (*p*:0.18; β [95% CI]:1.17 [−0.54; 2.88]; ρ:0.1149; E_point_:1.77; E_CI_: 1.00; Figure 2E). After birth, critical changes in lung and chest wall dynamics play a major role in developing breathing mechanics (31). With advancing PMA, lung alveoli mature both functionally and anatomically, improving lung-chest wall elasticity (32). We speculate that these changes play a major role in driving decreases in respiratory rate with advancing PMA. The contrasting results between apnoea and respiratory rate underlie the distinction between normal breathing and apnoea and further highlight the importance of utilising brain age gap as a biomarker for caffeine requirement in preterm infants.

Guidelines for discontinuing caffeine treatment are related to PMA, but there is variation in when treatment is stopped (33), depending on clinical discretion and guided by the infant’s physiological stability. Given that apnoea rate is related to brain age gap, we hypothesised that the age at which infants stopped caffeine treatment would be related to their brain age gap. In a subset of 27 infants who had their EEG recorded shortly before discontinuing caffeine treatment, we found that the age at which infants stopped caffeine treatment was correlated with their brain age gap (*p*:0.14; β [95% CI]:-0.26 [−0.61; 0.09]; ρ:-0.2839; E_point_:1.99; E_CI_:1.00; Figure 2**Figure 2**G) with infants with immature brain activity discontinuing caffeine at older ages (the decision to stop treatment was made by the clinical team who were unaware of the brain age gap or EEG of the infant; note all infants stop caffeine whilst in neonatal care, however, for most infants we did not record their EEG at the time at which they stopped caffeine treatment and so they were not included in this subset analysis). However, despite this association, some infants with relatively mature brain activity continued caffeine close to term-corrected age, and, *vice versa*, some infants with immature brain activity discontinued treatment at a much younger age. Using an infant’s brain age gap will provide an objective metric to identify when to discontinue treatment.

Finally, in an exploratory hypothesis-generating study in a subset of infants who had longitudinal recordings of vital signs throughout their hospital stay (see Methods), we investigated whether there was a relationship between brain age gap (recorded before infants discontinued caffeine), and apnoea rate in the 7 days after they discontinued caffeine treatment. Infants with more mature brain activity (defined as brain age gap >0) had 60% fewer apnoeas and 42% fewer oxygen desaturations after they stopped caffeine treatment compared with those infants with immature brain activity (brain age gap <0) (Apnoea rate, n=11: infants with mature brain activity: 0.41±0.40 apnoeas/hour [mean±SD]; infants with immature brain activity: 1.02±0.60; Figure 2H; Desaturations n=17: infants with mature brain activity: 0.26±0.38 desaturations/hour; infants with immature brain activity: 0.45±0.33; Figure 2I). As this exploratory study was limited in sample size and conducted to determine if there was any preliminary evidence of a relationship between brain age gap and apnoea rate after caffeine discontinuation, no statistical analyses were performed. This exploratory study is also limited in that it was a single-centre study comprising mainly Caucasian babies (Table 1). The study did not follow a pre-registered protocol and only examined the subset of infants that had longitudinal recordings at the time of stopping caffeine. These results should therefore be replicated in a prospective study before definitive conclusions can be drawn. Nevertheless, our findings provide promising initial evidence that physiological instability after discontinuation of caffeine is related to brain age gap.

The results presented here demonstrate that apnoea rate is related to brain age gap, not postmenstrual age, in infants from 31–36 weeks. We focused on this age range as this is the age at which doctors will consider whether or not infants require caffeine treatment, and unlike at younger ages, not all infants will experience apnoea. In infants of this age who are experiencing apnoea and/or desaturations, it can be difficult to disentangle the cause, which may be brain immaturity but also other factors such as infection or feeding problems – for which caffeine is not the best course of treatment. We propose that brain age gap (once further validated in other studies) could be used as an objective approach to dissociate the underlying mechanism of apnoea and determine whether it is appropriate to stop caffeine treatment. Future work should also assess whether EEG can be used as a biomarker for caffeine treatment in younger infants – whilst all extremely preterm infants are likely to need caffeine at birth, it is plausible that brain age gap may be a useful biomarker to indicate the dose of caffeine an individual will require.

We identified a significant relationship between apnoea rate and brain age gap, nevertheless, there was heterogeneity in responses. Within the statistical model, we adjusted for data length and the presence of infection, and additionally mode of ventilation (see Supplementary Results). We did not adjust for gestational age or postnatal age, as in our sample this was highly colinear with PMA (see Supplementary Results). Future work to investigate heterogeneity in responses is required. Finally, further work is needed to translate the results presented here into a clinically usable tool: EEG is already used in neonatal units, however, technology to incorporate brain age assessment within an EEG device is needed. Additionally, a prospective study evaluating the relationship between brain age and apnoea rate after stopping caffeine treatment is needed. This study could be used to build a model to identify the optimal brain age at which to stop caffeine treatment in premature infants, which could then be assessed in a randomised controlled trial.

In summary, we demonstrate that apnoea rate is related to brain age gap, not PMA, in infants from 31-36 weeks PMA. We also provide promising initial evidence that brain age gap is related to apnoea rate and oxygen desaturations in the week after caffeine is stopped. Whilst the utilisation of brain age as a biomarker for caffeine requirement requires additional validation, our current study provides a foundation for its use. There is a need for personalised medicine in neonatology; this study demonstrates that brain age gap is a key candidate biomarker for tailored caffeine treatment.

## Methods

### Study design and participants

This was a prospective cohort study conducted at the Newborn Care Unit of the John Radcliffe Hospital, Oxford University Hospitals NHS Foundation Trust, Oxford, UK between 2019 and 2023. Clinically stable preterm infants aged between 31 and 36 weeks postmenstrual age (PMA) at the time of test occasion were eligible to be included in the study (from an age of 31 weeks and 0 days up to 36 weeks and 6 days). Infants were excluded if they had grade III or IV intraventricular haemorrhage, hypoxic ischaemic encephalopathy, congenital malformations, were on mechanical ventilation, receiving opioid analgesics at the time of study, or if there was a history of maternal substance misuse during pregnancy. These conditions are known to impact brain activity and may interfere with the interpretation of the EEG by acting as confounders. Previous studies on brain age models utilised similar inclusion and exclusion criteria (17,18).

The study was approved by the National Research Ethics Service (references: 19/LO/1085, 12/SC/0447). Eligible families were given verbal and written information about the study, and written parental consent was signed before inclusion in the study. The study conformed to the standards set by the Declaration of Helsinki and Good Clinical Practice.

An initial total of 76 infants were studied on 141 separate test occasions; 53 infants were included in one to three test occasions, 23 infants took part in the ongoing *Breathing and Brain Development* study (34) where their EEG is recorded approximately once a week and their vital signs recorded continuously whilst on the Newborn Care Unit (included on average on 3±1 [mean ± SD] occasions). During each test occasion (which aimed to last ∼2 hours), simultaneous electroencephalography (EEG) and vital signs (respiration, heart rate and oxygen saturation) were recorded. Three test occasions were excluded due to very short recordings of inter-breath intervals (see ‘computing respiratory and apnoea rate’), leaving a total of 74 infants studied on 138 test occasions included in the analysis.

### Data acquisition

#### Vital signs recordings

Each infant had continuous vital signs monitoring as the standard of care. Vital signs were monitored using Phillips IntelliVue MX800 or MX750 monitors and data were continuously downloaded from the vital signs monitor using an electronic data capture software (iXtrend, ixitos, Germany). For infants in the *Breathing and Brain Development* study, vital signs were downloaded continuously throughout the infant’s hospital stay; the recordings from the day of the EEG test occasion are included in the main analysis (i.e., up to 24 hours of recording). For other infants, vital signs were recorded during EEG data acquisition for an average of 1.5 hours (median, range: 0.4-3.3 hours).

Heart rate, oxygen saturation and respiratory rate (calculated by the monitor) were downloaded onto the laptop at a sampling rate of 0.97 Hz; the electrocardiograph (ECG, to measure heart rate) at 250 Hz and recorded with three electrodes placed on the infant’s chest; the impedance pneumography (IP, to measure respiration) at 62.5 Hz and recorded from the chest electrodes, and the photoplethysmography (PPG, to measure oxygen saturation and pulse) at 125 Hz from a probe placed on the infant’s foot or hand.

#### EEG recordings

EEG was recorded from DC to 400 Hz using a SynAmps RT 64-channel headbox and amplifiers and CURRYscan7 neuroimaging suite (Compumedics Neuroscan) at a sampling rate of 2 kHz. Eight electrodes were placed at Cz, CPz, C3, C4, Oz, FCz, T3, and T4, according to the modified international 10-20 system. The ground electrode was placed at FPz and the reference electrode at Fz. Infant scalp preparation was done with NuPrep gel (D.O. Weaver and Co., Aurora, USA) to achieve impedances of 5-10 kΩ. A conductive paste (Elefix EEG paste, Nihon Kohden, Tokyo, Japan) was used to attach disposable Ag/AgCl cup EEG electrodes (Ambu Neuroline, Ballerup, Denmark) to the infant’s scalp. At the beginning of each recording, approximately 10 background annotations were time-locked on the EEG and vital signs recordings by manually pushing a button connected to a bespoke triggering device (the PiNe box) (35) which simultaneously annotates the EEG and vital signs recordings. This enabled synchronisation of the vital signs and EEG recordings. Background annotations were performed by a researcher during a period when the infant was quietly resting in their cot/incubator. Resting-state EEG was recorded for an average of 1.7 ± 0.7 hours (mean ± SD). No clinical records on the normality of the EEG were available.

#### Tactile and visual stimuli

A tendon hammer was used to lightly tap the infant’s foot to elicit touch responses. At least 10 gentle touch stimuli, with an inter-stimulus interval of approximately 10 seconds (longer if the infant was unsettled) were applied. The tendon hammer was modified with a built-in force transducer (Brüel & Kjær, Denmark) that sends a trigger pulse at the point of stimulation (36). For visual-evoked responses, the stimulus was a flash of light presented using a Lifelines Photic Stimulator (intensity level 4, approximately 514 lumens). The Photic Stimulator was placed approximately 30 cm away from the infants, in line with their direct field of vision and a train of 10 light flashes was delivered at least 10 seconds apart. The tactile and visual stimuli (order pseudo-randomised) were performed either at the start or end of the EEG recording. The stimuli were annotated simultaneously on the EEG and vital signs recordings using our custom-made time-locking device (the PiNe box) (35).

#### Recording of infant demographics and clinical details

Relevant demographic and clinical information were obtained from the medical notes of each infant and updated at each test occasion where relevant. These included the infant’s age at birth and sex, and at the time of the test occasion, the postmenstrual age, current mode of ventilation and presence of infection. Infection was defined based on the neonatal guidelines for infection treatment at the John Radcliffe Hospital as either an elevated C-reactive protein of >10mg/L, abnormal white blood cell count (>20 x 10^9^/L or <5 x 10^9^/L), a positive blood culture; or a symptomatic infant with predisposing factors for sepsis receiving antibiotics at the time of EEG recording.

### Data analysis

All analyses were performed in MATLAB (ver. 2022b; MathWorks Inc., Natick, USA). In brief, we first computed respiratory and apnoea rates from the impedance pneumography (IP) signals, and brain age gap from 20 minutes of resting state EEG activity and sensory-evoked (visual and tactile) potentials. Next, we constructed linear (mixed-effects) models to test if the respiratory outcomes – respiratory and apnoea rates – were associated with either PMA or brain age gap. We also explored whether the PMA at which caffeine treatment is discontinued relates to brain age gap. In a final exploratory analysis to explore whether the brain age gap could be utilised as a predictive biomarker for stopping caffeine treatment, we investigated whether the rate of episodes of physiological instabilities (apnoea and oxygen desaturation rate) in the week after caffeine was stopped is related to brain age gap. Each of these analysis steps are outlined in more detail below.

#### Computing respiratory and apnoea rate

To obtain respiratory and apnoea rates, IP signals were processed using an algorithm validated for the identification of inter-breath intervals and apnoeas in infants (37). Briefly, the algorithm first filters the signal to reduce the noise introduced by movements and cardiac activity. An adaptive amplitude threshold (set at 0.4 times the standard deviation of the signal over the preceding 15 breaths) is used to identify individual breaths. Finally, a support vector machine algorithm was applied to all potential episodes of apnoea (defined as inter-breath intervals longer than 15 seconds) to remove periods of low amplitude erroneously identified as apnoeas due to noise or shallow breathing. This model was trained and validated on IP signals acquired during a clinical trial (38). The data from the clinical trial was completely independent from our current data sample. Long inter-breath intervals (IBI) marked as noise/shallow breathing were discarded from further analysis. We then computed respiratory and apnoea rates estimated from all available IBIs on the day of the EEG recording, which was 8.5 ± 8.7 hours (mean ± SD over recordings). We computed respiratory rates (expressed in times per minute) and apnoea rates (in times per hour) estimated from all available IBIs on the day of the EEG recording. We excluded three recordings which had fewer than 100 IBIs available. Apnoea rate was calculated as the number of apnoeas (IBIs greater than 15 seconds) per hour. Respiratory rate was computed as the median number of breaths per minute (i.e., 60 divided by the median IBI of the recording).

#### Estimating brain age gap from EEG activity

To obtain the brain age gap, we first computed brain age using two brain age models (17,18). These two models estimate the functional maturation of the brain using resting state (18) and sensory-evoked EEG activity (17), respectively. These models aim to capture different developmental aspects. The resting state model is an index for the maturation of ongoing brain activity and the sensory-evoked model for the maturation of sensory pathways (Supplementary Figure S1). Whilst both models use features extracted from different neural systems, all features change in an age-related manner. Both are machine learning models which are trained to estimate the age of the infant from the EEG activity, with the aim of such models being to identify infants who deviate from normal neurodevelopment. Thus, brain age is correlated with PMA, and large differences between PMA and brain age may be indicative of abnormal outcome (17,18). Brain age gap was defined as the mean brain age of the two models minus PMA (note, brain age gap has also been referred to in the literature as the brain age delta). Figure S2 compares brain age and respiration/apnoea rate.

Before estimating the brain age gap, EEG data were first pre-processed using the Brainstorm (version 3) (39) and EEGLAB (version 2022.1) (40) toolboxes. EEG time series were filtered with pass-band edges at 0.1 and 30 Hz by consecutively high-pass (Hamming windowed-sinc FIR filter with a cut-off frequency at 0.05 Hz) and low-pass filtering the signals (Hamming windowed-sinc FIR filter with a cut-off frequency at 33.75 Hz).

To calculate the infant’s brain age from their sensory-evoked responses (visual and tactile), we used the model developed by Zandvoort and colleagues (2024) (17). Briefly, this model capitalises on the PMA-related changes in the morphology of sensory-evoked potentials during the preterm period. The sensory-evoked data were first epoched from - 0.5 to 1.0 seconds around the stimulus onsets for channels Oz and Cz. Epochs were baseline normalised by subtracting the mean amplitude of the time window between −0.5 and 0 seconds. Epochs were then visually inspected, and those with excessive amplitudes or artefacts were excluded (an average of 10% of epochs were excluded). Individual test occasions with fewer than five epochs of sensory-evoked data remaining after artefact rejection were excluded from sensory-evoked brain age estimation (9 out of 138 recordings), and brain age was estimated using the resting state model alone for these recordings. Visual and tactile evoked potentials were fitted to neurodynamic response functions based on the characteristic waveforms of the evoked stimulus responses (17) (Figures S3-S4 show the average evoked potentials). Zandvoort et al. (2024) extracted these characteristic waveforms using principal component analysis. The visual and tactile responses at channels Oz and Cz, respectively, can be captured by four visual and two tactile neurodynamic response functions (17). Before fitting, evoked responses were Woody filtered to the neurodynamic response functions to improve their temporal alignment, allowing for individual infant differences in the latency of the evoked response (maximum jitter: 50 ms; number of iterations: 1) (41). Neurodynamic response functions were then fitted to the individual’s evoked responses using a linear regression. The regression’s slope indicates the magnitude of the evoked response. The slopes were obtained for the six neurodynamic response functions and forwarded as input for the sensory brain-age model, which consisted of a support vector regression with a linear kernel function using MATLAB’s fitrsvm function (version 2022b; MathWorks). The output of the model is the predicted sensory brain age for the infant (for further details, see Zandvoort et al., 2024 (17)). The mean absolute error for data presented in this study is comparable to the data presented in the original paper (Supplementary Figure S5).

To calculate the infants’ resting state brain age, we used the model developed by Ansari and colleagues (2024) (18). Data were first downsampled to 64 Hz, and the first 20 minutes of every recording was selected for analysis as this should warrant reliable brain age estimates (18). EEG recordings were started when the infant was settled in their cot/incubator and no overt artefacts were present (data length: 105 ± 41 minutes [31, 193]; mean ± standard deviation [range]). Since our studies comprised resting-state and stimulus-evoked recordings, we selected a 20-minute window in which no visual or tactile stimuli were applied. This enabled us to standardise the recording length used for analysis across infants. However, in line with Ansari et al. (2024), we show that brain age predictions estimated over 20 minutes are comparable to predictions from the full recordings (Figure S6A) and that using the full recording to estimate the brain age does not alter the relationship between apnoea rate and brain age gap (Figure S6B). For one recording, the total data duration was less than 20 minutes, meaning that we did not have sufficient data to calculate a reliable resting state brain age. For this recording, we only used the brain-age estimate of the sensory model. For all recordings, the bipolar derivative between channels C3 and C4 was used for brain age estimation. The data were input into the deep neural network, and the output of the model was the predicted resting state brain age for the recording (for further details, see Ansari et al. 2024 (18)). Briefly, this algorithm segments the data into 30-second epochs and estimates a brain age for each of them for ten iterations (the model consisted of a 10-learner ensemble method). To obtain a single brain age value, the median over all ten ensembles was taken for every 30-second epoch, after which a median over all 30-second epochs resulted in a single value. To validate the output, Ansari et al. (2024) also used an independent test set and found a MAE of 1.03 weeks for the held-out dataset. Their model also yielded a significantly lower MAE than a null model estimating the mean PMA of the dataset. Model applications revealed mean absolute errors of 1.22 and 0.77 weeks for the sensory-evoked and resting state models within the data studied here. These accuracies are comparable to those reported in the original studies which were 1.41 and 0.79 weeks. Moreover, for model training, Ansari et al. (2024) used data of infants with normal neurodevelopmental outcome at 24-month follow-up (assessed with BSID-II). Their model was validated on EEG data that were labelled as normal by a neurophysiologist. Neither brain age models take into account the different sleep states. Whilst it is known that sleep states alter EEG activity in infants (21), derived brain ages are marginally affected by it according to earlier versions of the model (42).

We obtained a single brain age prediction by averaging the brain ages of both models. Since the models may estimate brain age with systematic prediction errors relative to PMA (16), we corrected for this by first fitting a linear model between the combined brain age and PMA. We next predicted PMA from this model and defined the model bias as the difference between the PMA and predicted PMA (meaning that brain age gap and PMA are completely uncorrelated). Next, brain age was corrected by subtracting the bias from the brain age (relative to PMA). This age-bias correction is a standard approach in the brain age field (16,43). This also ensured that the brain age gap and PMA were uncorrelated. However, without this bias correction, similar results were obtained (Supplementary Figure S7).

#### Probability density functions for the inter-breath intervals

To study how the IBI dynamics changed with PMA and brain age gap, we created probability density functions. To do so, we first created a frequency distribution from the IBIs of each recording a resolution of 0.1 seconds. IBIs longer than 50 seconds were removed from further analysis. Distributions of each recording were normalised so that the area under the curve was equal to 1, allowing for inter-recording comparisons. We next transformed these recording-specific probability density functions into functions specific for a PMA/brain age gap. For this, a weight of 1 or lower was assigned to the recording-specific probabilities, depending on the infant’s PMA/brain age gap. Weights were determined using a Gaussian window with a full width at half maximum of 13 days. Weightings for differences of more than 14 days were set to 0. Distributions that were specific to a certain PMA/brain age gap were computed as the weighted median of all distributions together with their weights (44). We created probability density functions between 31 and 36 weeks for the PMA and between −3 and 3 weeks for the brain age gap.

#### Statistical analysis

To compare PMA and brain age gap with apnoea rate and respiration rate, we used linear mixed-effects models (computing four models for each of apnoea rate and respiration rate against PMA and brain age gap separately). Previous literature suggested that experiencing apnoeas is inversely associated with gestational age at birth (27,30). The course of this relationship is, however, unknown which is why we used linear models as a starting point. Nevertheless, we tested the assumption of linearity (see Supplementary material and Table S1) using the mfp package in R. Moreover, it is not known how apnoea rate varies with brain age gap. A linear fit was appropriate for the data collected here so as not to overfit the data; however, whether a linear association underlies the brain age gap/apnoea relationship needs to be studied in future research. Data length of the IBI recordings and infection (‘no’, ‘suspected’, or ‘treated’) were included as fixed factors and infant was included as a random factor (to account for the repeated test occasions in the same infant). Data length and infection were included as confounding factors. P-values (for the slopes of PMA and brain age gap) and partial correlation coefficients and their 95% confidence intervals are reported from these models. The normality assumption was verified with Q-Q plots. Further discussion of the fixed factors to include within the model, and assessment of the results with alternative factors is included in the Supplementary Material. To test for the robustness of the associations and to estimate the strength of unmeasured confounders, we report E-values (45). Reporting E-values is the most common method for observational medical studies (46). We provide E-values for the observed association estimate and the confidence interval limit that is closest to null, computed via https://www.evalue-calculator.com. For visualisation, (we used the plotAdjustedResponse function in MATLAB to adjust for the fixed factors (calculating a separate model without the random effect of infant to do so). Regression lines are visualised by calculating the line-of-best-fit on the adjusted data.

To test if apnoea rate is better associated with brain age gap than PMA, we first bootstrapped the partial correlation coefficient between apnoea rate and PMA using 10,000 repetitions. This allowed us to generate a 95% confidence interval for the association between apnoea rate and PMA. To test if the partial correlation coefficient between brain age gap and apnoea rate lies outside this interval, the significance value was defined as the number of bootstrapped partial correlation coefficients that were equal to or lower than the true partial correlation coefficient between brain age gap and apnoea rate. Significance was tested one-tailed with an alpha level of 0.05.

For the association between brain age gap and age at caffeine termination, we focused on a subset of 27 infants who had their brain age gap assessed within two weeks before they stopped caffeine treatment (note that infants were all receiving caffeine during the EEG recording). Note, the age at which caffeine was stopped was dependent on clinical decision and independent of the research study. A linear regression was used with infection status as a fixed factor (data length was not included as a fixed factor as all EEG was analysed using a fixed data length, and respiratory data was not analysed for this question). For visualisation, we used the plotAdjustedResponse function adjusting for the fixed effect of infection status only and the regression line was calculated as the line-of-best-fit of the adjusted data.

In the exploratory analysis, we compared brain age gap before infants stopped caffeine treatment with apnoea rate and oxygen desaturation rate in the week after they stopped caffeine treatment. In a subset of infants (n=17) with longitudinal recordings of vital signs throughout their hospital stay, infants were divided into two groups depending on their brain age gap – either mature if their brain age gap >0 weeks or immature if their brain age gap <0 weeks. The brain age gap was calculated from a single EEG recording taken in the two weeks before they stopped caffeine. Apnoeas and desaturations were identified from the recordings in the 7 days after caffeine was stopped (the age at which caffeine was stopped was dependent on clinical decision and independent of the research study). Desaturation events were defined as 10-second periods in which the oxygen saturation was less than 80% (38). The type of vital signs monitoring was dependent on clinical decision, and so 6 out of 17 infants did not have their IP signal measured during this week (only oxygen saturation and pulse were monitored using a PPG), and they were consequently excluded from the apnoea rate analysis. Due to the limited sample sizes, and as this was an exploratory hypothesis-generating study, we did not apply any statistics and reported outcomes as mean and standard deviation.

## Acknowledgements

This work is funded by the Wellcome Trust and Royal Society through a Sir Henry Dale Fellowship awarded to CH (grant number: 213486/Z/18/Z). FU is funded by the Commonwealth Scholarship Commission. The funders had no role in study design, data collection and analysis, decision to publish, or preparation of the manuscript. We would like to thank the babies and their parents who took part in this research.

## Data and code availability

Example resting state data as well as processed data (including brain age gap, apnoea rates and predictor variables for each individual) and the codes for the analysis are available at: https://github.com/CoenZandvoort/Brain_maturity_respiration. The brain age models can be found at: https://github.com/amirans65/brainagemodel and https://gitlab.com/paediatric_neuroimaging/sensory-brain-age-model. The algorithm for inter-breath interval detection is online at: https://gitlab.com/paediatric_neuroimaging/identify_ibi_from_ip. Due to ethical requirements, the raw data that support the findings of this study are available upon request to the corresponding author.

## Supplementary Material

### Supplementary Results

#### Regression models

This section contains some further details on the linear models used in the analysis. We considered a number of factors within the statistical analysis and expand on this further here.

Potential factors to include in the model were considered to be:

1. Data length of the impedance pneumography (IP) recording – the recording used to calculate apnoea rate and respiratory rate. Note that data length used in the EEG analysis was the same for all recordings and so was not considered as a factor.
2. Presence of infection
3. Current mode of ventilation
4. Gestational age at birth
5. Postnatal age

We discuss each of the factors in turn in the following.

1. Data length of IP signal. Data length varied substantially across studies due to the inclusion of two study designs – some infants were part of the *Breathing and Brain Development* study and so had up to 24 hours of data from the day of EEG recording, other infants were included for a shorter (approximately 1-2 hour) recording (see Methods). As apnoeas are a relatively rare event – occurring approximately once an hour – apnoea detection using short recordings is at risk of underestimating apnoea rate, and so, data length was included in the statistical model of apnoea rate. Respiratory rate (the mean respiration during normal breathing) can be accurately estimated with short data lengths and so data length was not included in models of respiratory rate.
2. Presence of infection was considered a confound variable and so was included in the main statistical model. Given that the presence of infection may increase apnoea rate, we further validated the main results as presented in Figure 2, by also considering the subset of infants who did not have infection at the time of study (Figure S10). In line with our main results, apnoea rate was significantly related to brain age gap in this subset (*p*: 0.02; β [95% CI]: −0.21 [−0.38; −0.03]; ρ: −0.21) but not postmenstrual age (PMA) (*p*: 0.19; β [95% CI]: −0.07 [−0.18; 0.04]; ρ: −0.12).
3. Mode of ventilation is related to PMA, with younger infants more likely to be receiving high flow therapy and older infants more likely to be self-ventilating. All infants in this study were receiving either high flow therapy, low flow therapy or self-ventilating at the time of study. Apnoea rate and respiratory rate are also related to mode of ventilation and so mode of ventilation may act as a mediator variable (rather than a confound variable). We therefore chose not to include it in the main statistical analysis but have also assessed the results with inclusion of this factor (Figure S11). Apnoea rate reveals a strong trend with brain age gap, but not with PMA (brain age gap: *p*: 0.06; β [95% CI]: −0.18 [−0.37; 0.01]; ρ: −0.16; PMA: *p*: 0.67; β [95% CI]: −0.03 [−0.15; 0.10]; ρ: −0.04). Moreover, to further validate the main results, we conducted a subgroup analysis with self-ventilating infants (Figure S). This demonstrated a correlation in the same direction between apnoea rate and brain age gap (*p*: 0.10; β [95% CI]: −0.20 [−0.44; 0.04]; ρ: −0.17; PMA: *p*: 0.51; β [95% CI]: 0.04 [−0.09; 0.18]; ρ: −0.07) but due to the lower sample size in this group (n=57 infants on 89 occasions) we were likely underpowered to demonstrate a significant effect.

We considered gestational age (age in weeks at birth) and postnatal age (expressed as age in weeks after birth). Both are highly correlated with PMA (age in weeks at the time of study, which is the sum of gestational age and postnatal age) (Figure S13) and we, hence, did not add these to the model due to issues with collinearity.

##### Linearity of factors

We tested the assumption of linearity for the linear models (Figure 2A, B, D, E, and G) using multivariable fractional polynomial models. These models determine whether relationships are better modelled through non-linear relationships by finding the most appropriate functional form for every continuous predictor. Using the mfp package in R, we estimated these most appropriate forms and statistically tested them against the factor being included as linear. We included the same factors as presented in the main text (Figure 2A, B, D, E, and G; i.e., infection and data length when estimating apnoea rate and infection for respiration rate). For all models, infant was added as random factor. The p-values of Table S1 represents the difference in deviances between the fractional polynomial and linear models. As indicated by this table, none of the p-values for any model were statistically significant, confirming the linearity assumption.

**Table S1.**
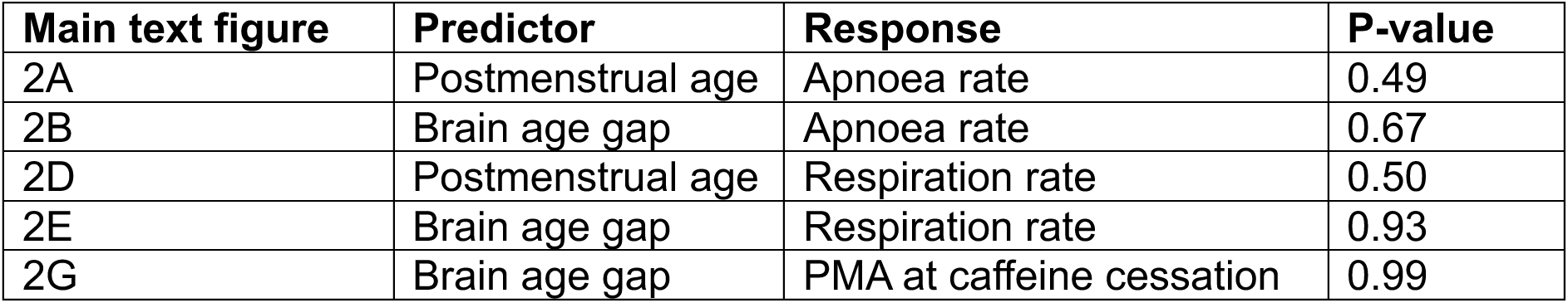
Statistical output of fractional polynomials relative to linear models.

#### Exploratory hypothesis-generating findings

In the main text we identified a possible relationship between brain age gap and apnoea rate in the week after caffeine treatment is discontinued, in an exploratory hypothesis-generating study. For a power calculation for future studies, based on the apnoea rates from our analyses, we obtained an effect size of 1.19 using these values in G*power 3.1, and together with an α of 0.05 and power of 0.95, the required sample size is *n* = 34 for both groups (i.e., 17 participants per group).

---

**Figure S1.**
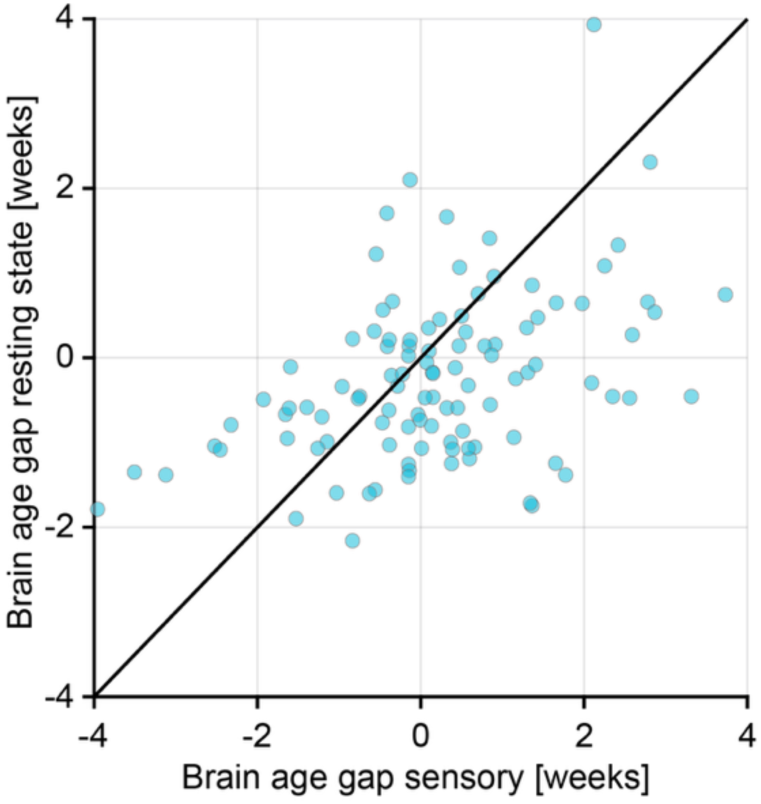
Brain age gap estimates from the two brain age models (computed from the sensory-evoked and resting state EEG activity). Brain age gap is computed relative to the age of the infant, with negative values indicating that the brain activity is immature relative to the infant’s PMA, and positive values indicating that the brain activity is more mature relative to the infant’s PMA. Each dot indicates an individual test occasion.

**Figure S2.**
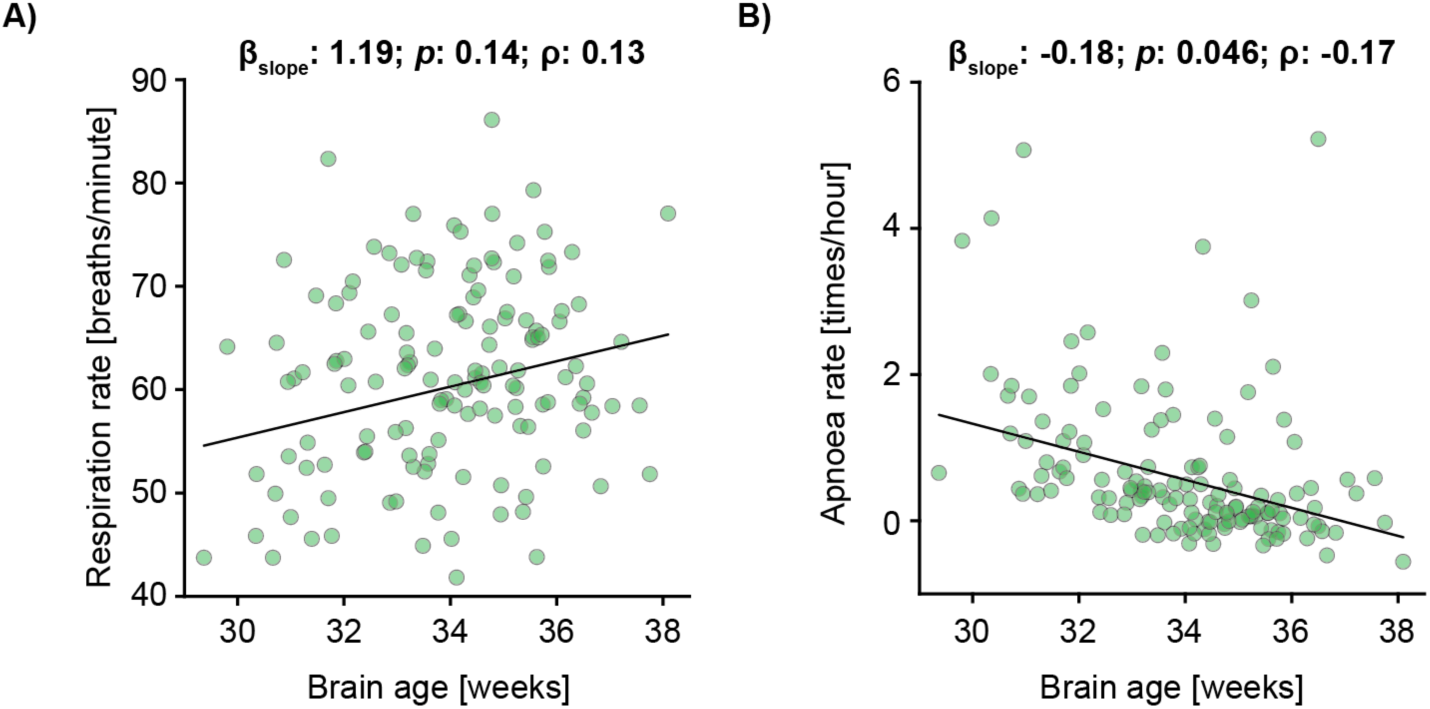
Respiration and apnoea rate compared with brain age. Brain age associations with **A)** respiration rate and **B)** apnoea rate. Brain age gap (as presented in the rest of the text) is defined as the difference in brain age minus postmenstrual age (PMA) (i.e. brain age is PMA plus brain age gap). Each dot indicates an individual test occasion (74 infants were studied on 138 test occasions). Black line is the mean fit of the linear model. Regression models are adjusted for infection, data length of inter-breath intervals, and postmenstrual age.

**Figure S3.**
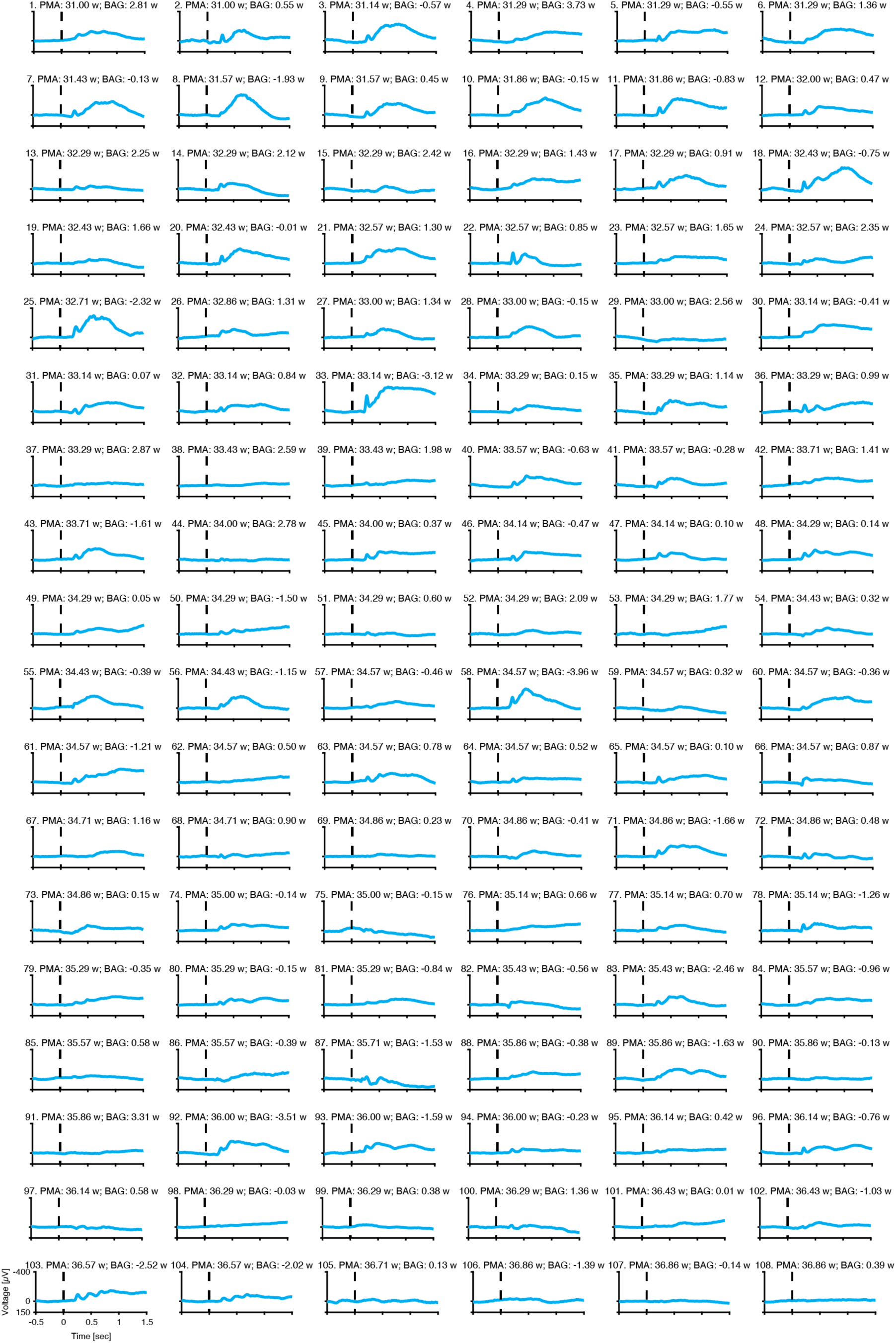
Recording-specific evoked potentials for the visual stimuli. Average (across all visual stimuli) evoked response for each individual recording, with the infants age and brain age gap indicated in the title of each subplot. Recordings are ordered according to increasing postmenstrual age (PMA). Stimulus was applied at Time = 0 sec. BAG: brain age gap; w: weeks.

**Figure S4.**
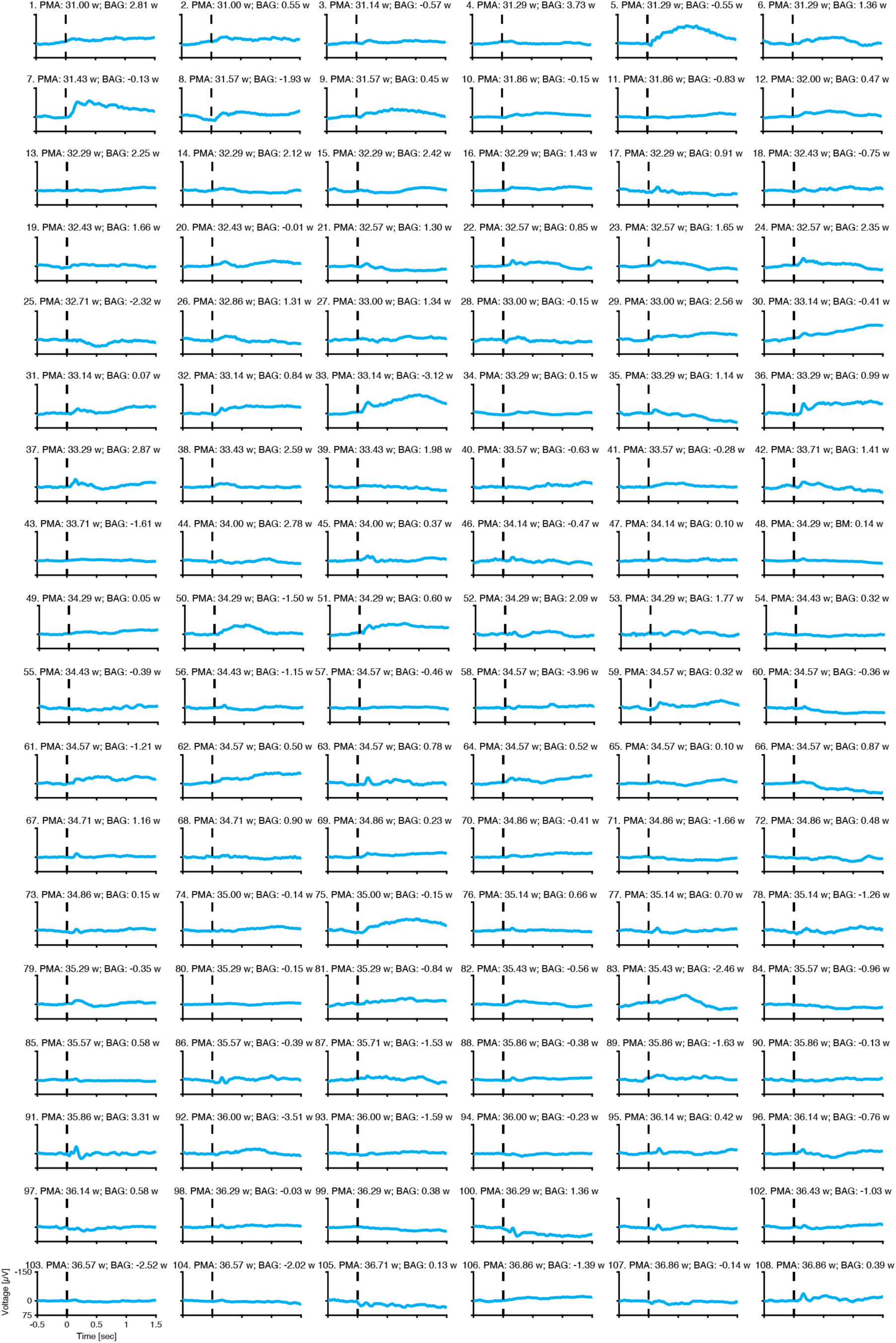
Recording-specific evoked potentials for the tactile stimuli. Average (across all visual stimuli) evoked response for each individual recording, with the infants age and brain age gap indicated in the title of each subplot. Recordings are ordered according to increasing postmenstrual age (PMA). Stimulus was applied at Time = 0 sec. BAG: brain age gap; w: weeks.

**Figure S5.**
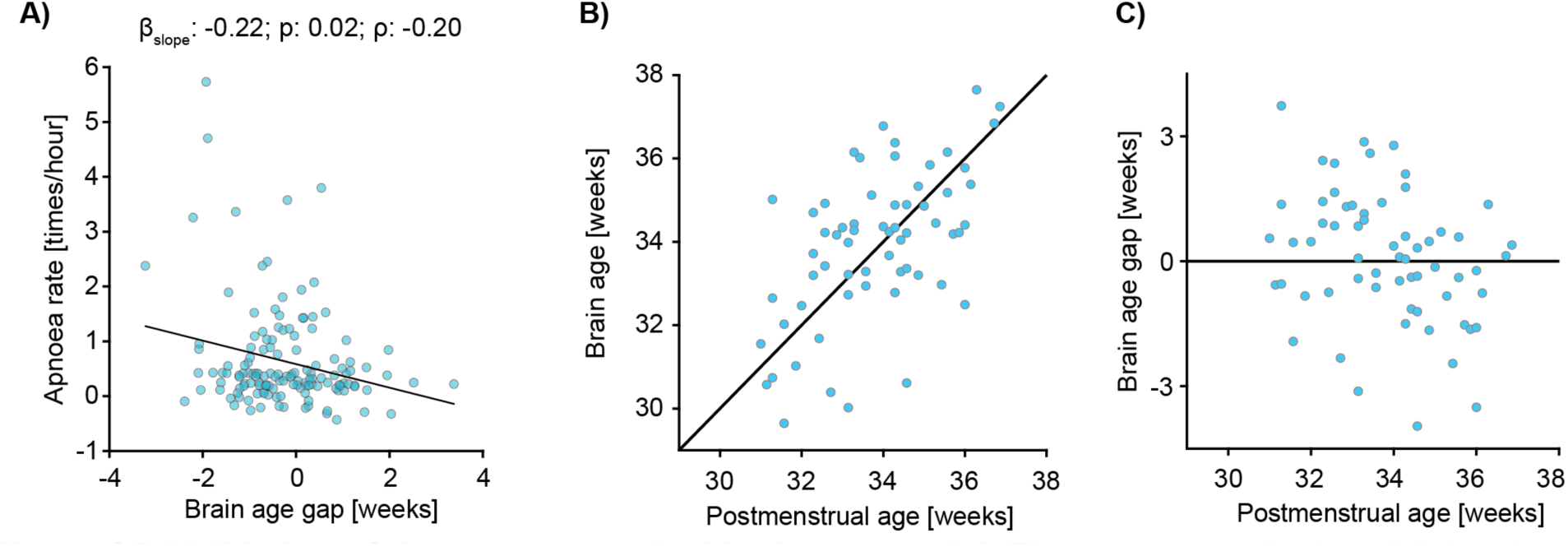
Validation of the sensory-evoked brain age model. The sensory-evoked model developed by Zandvoort et al. had an overlap with the data presented in this paper of 59 [out of 128] recordings from 48 babies. To check that this did not affect the results presented in the main text we investigated **A)** the relationship between brain age gap and apnoea rate excluding the sensory brain age estimates of babies that were part of the training set in Zandvoort et al. This showed similar results. Further, we validated the sensory-evoked model on the data that was not part of the original training set. **B-C**) Sensory brain age (gap) relationships with postmenstrual age for the data that was not included in the training set. The mean absolute error for this data was 1.22 weeks which is comparable to the mean absolute error of 1.41 weeks for the training dataset in the original paper.

**Figure S6.**
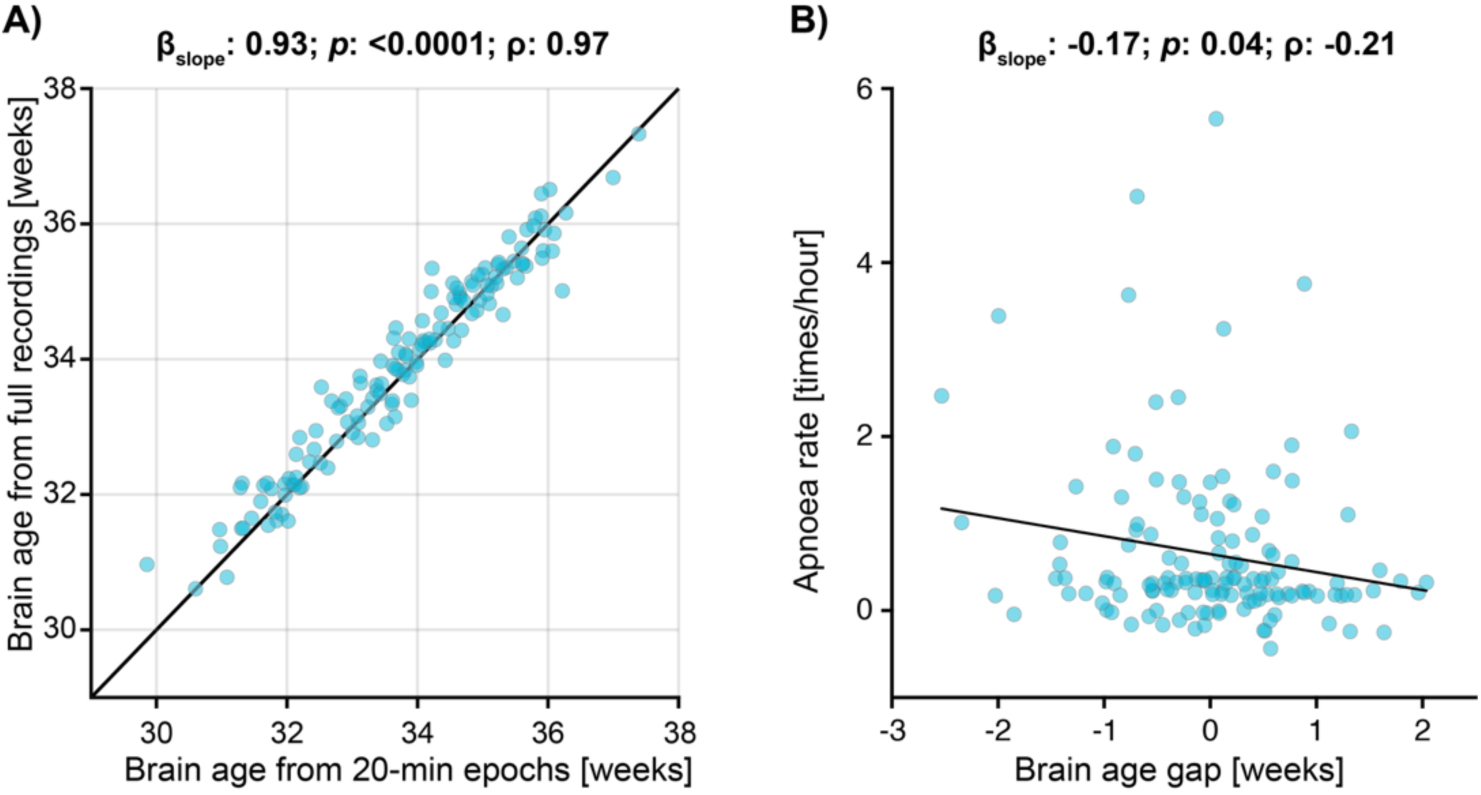
Effects of data length on resting state brain ages. **A)** Resting state brain ages predicted by 20-minutes epochs against those estimated over full recordings. The 20-minute epochs were used in the main results as this provided a consistent data length across all recordings and was the minimum data length which can be reliably used by the model as identified by Ansari et al.^14^ **B)** Apnoea rate association with brain age gap when full recordings are used to estimate resting state brain age. The black line is the mean fit of a linear mixed effects model. Brain age gap is computed relative to the age of the infant, with negative values indicating that the brain activity is immature relative to the infant’s PMA, and positive values indicating that the brain activity is more mature relative to the infant’s PMA. For both panels, each dot indicates an individual test occasion.

**Figure S7.**
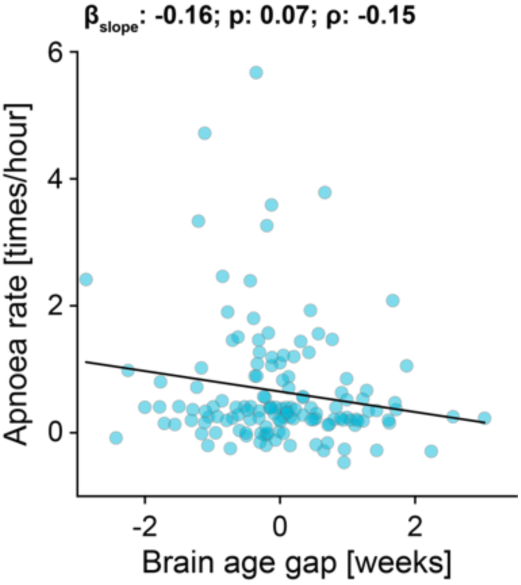
Effects of bias correction on resulting association between apnoea rate and brain age gap. Brain age gap is estimated from both resting state and sensory model. However, here we did not correct for deviations between brain age and PMA, which allowed for correlations between brain age gap and PMA. This still resulted in a significant relationship between brain age gap and apnoea rate. The black graph is the mean fit of a linear mixed effects model. Brain age gap is computed relative to the age of the infant, with negative values indicating that the brain activity is immature relative to the infant’s PMA, and positive values indicating that the brain activity is more mature relative to the infant’s PMA. Each dot indicates an individual test occasion.

**Figure S8.**
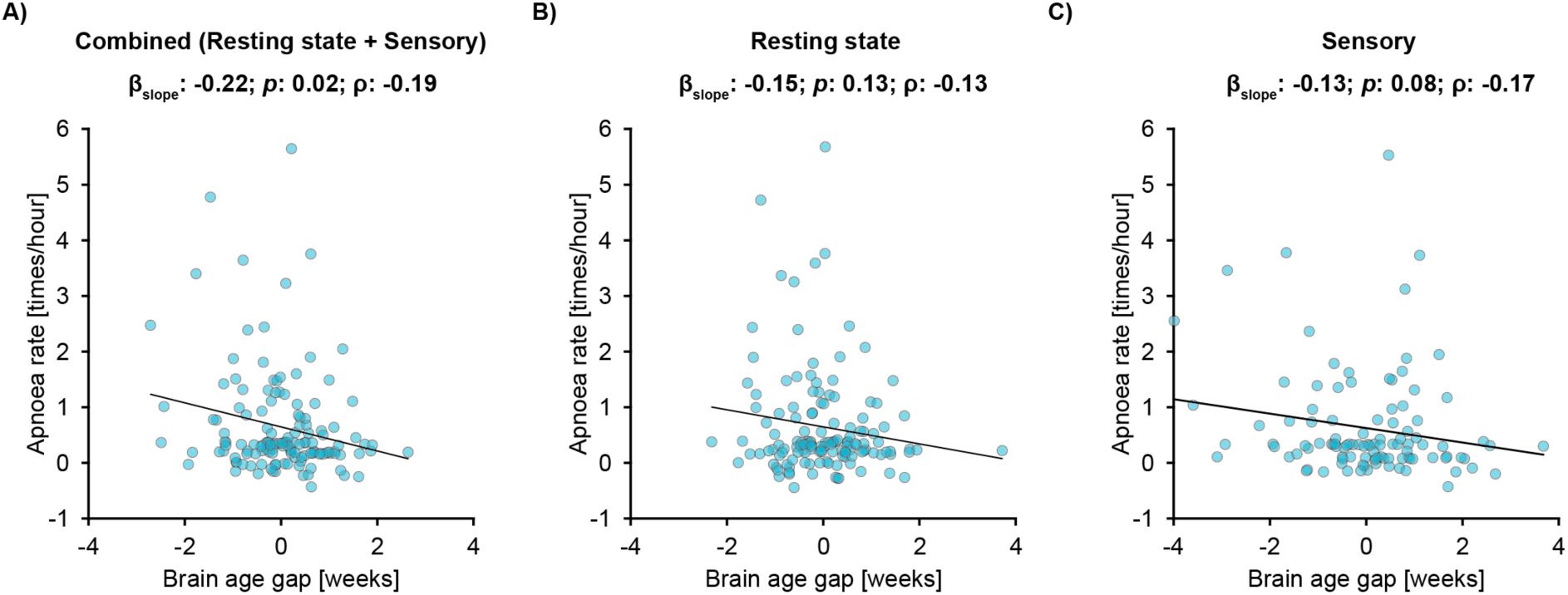
Apnoea rate against brain age gap using the brain ages from different models. Apnoea rate associations with **A)** both models, **B)** resting state model, and **C)** sensory model. Brain age gap is computed relative to the age of the infant, with negative values indicating that the brain activity is immature relative to the infant’s PMA, and positive values indicating that the brain activity is more mature relative to the infant’s PMA. Each dot indicates an individual test occasion. Black line is the mean fit of the linear model. Regression models are adjusted for infection and data length of inter-breath intervals.

**Figure S9.**
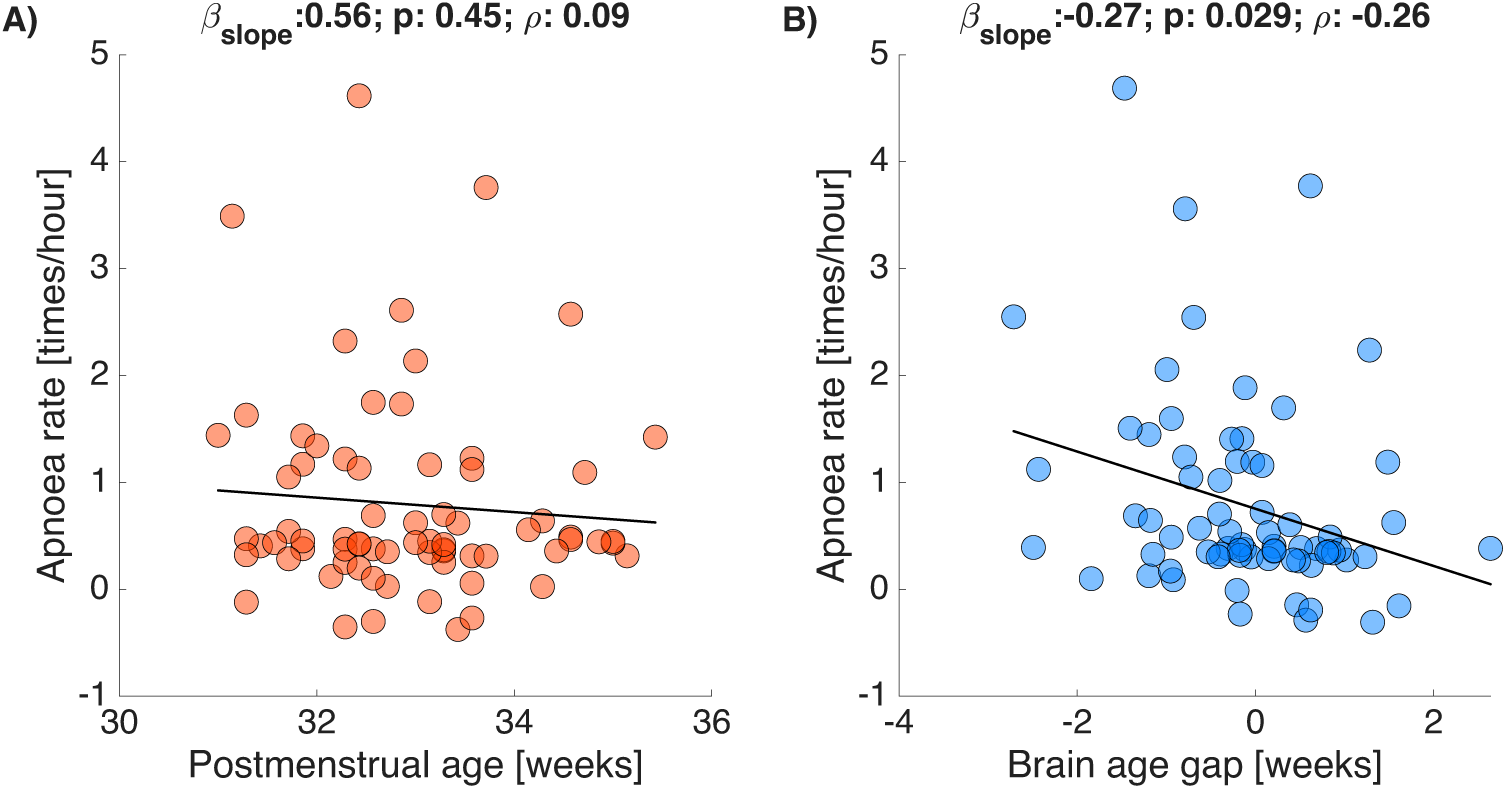
Apnoea rate against postmenstrual age and brain age gap for infants receiving caffeine at the time of study. Apnoea rate associations with **A)** postmenstrual age (PMA) and **B)** brain age gap. Brain age gap is computed relative to the age of the infant, with negative values indicating that the brain activity is immature relative to the infant’s PMA, and positive values indicating that the brain activity is more mature relative to the infant’s PMA. Each dot indicates an individual test occasion (40 infants were studied on 71 test occasions). Black line is the mean fit of the linear model. Regression models are adjusted for infection and data length of inter-breath intervals.

**Figure S10.**
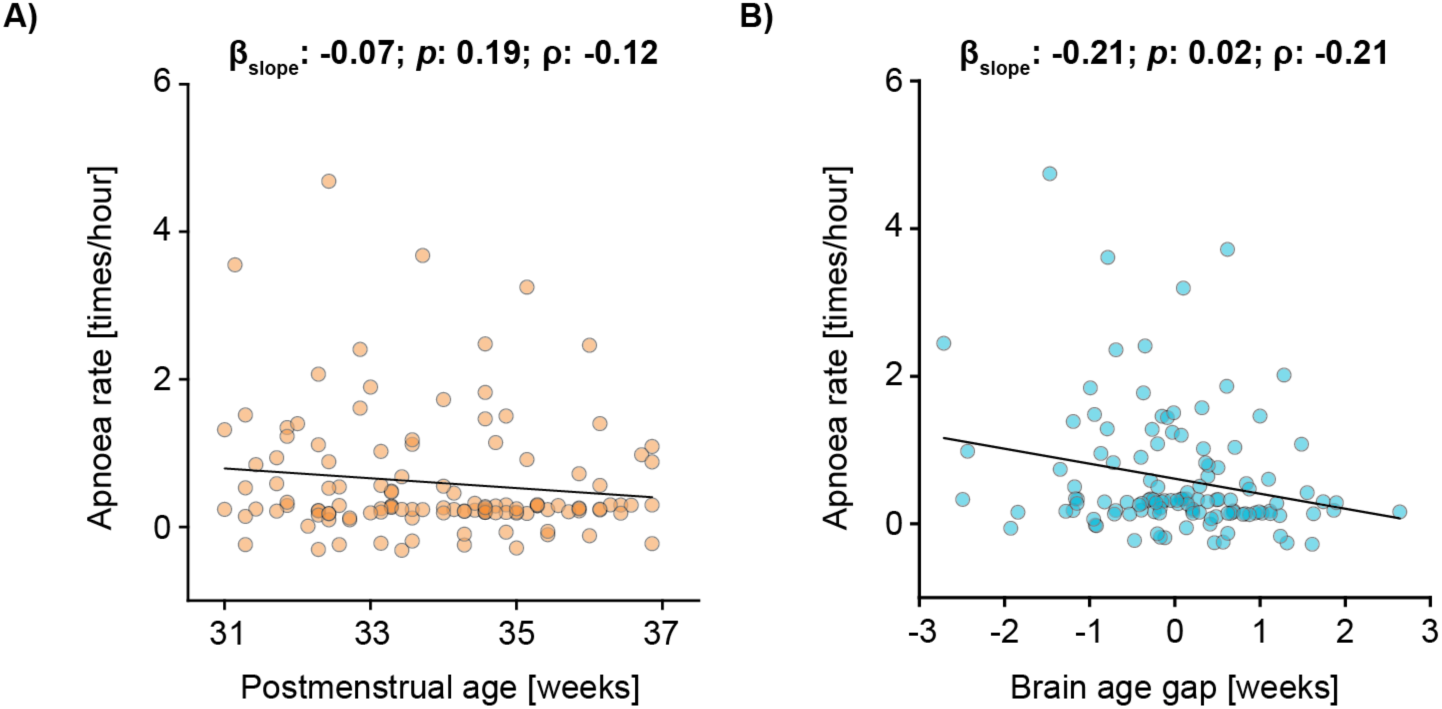
Apnoea rate against postmenstrual age and brain age gap for infants without infection at the time of study. Apnoea rate associations with **A)** postmenstrual age (PMA) and **B)** brain age gap. Brain age gap is computed relative to the age of the infant, with negative values indicating that the brain activity is immature relative to the infant’s PMA, and positive values indicating that the brain activity is more mature relative to the infant’s PMA. Each dot indicates an individual test occasion (69 infants were studied on 121 test occasions). Black line is the mean fit of the linear model. Regression models are adjusted for infection and data length of inter-breath intervals.

**Figure S11.**
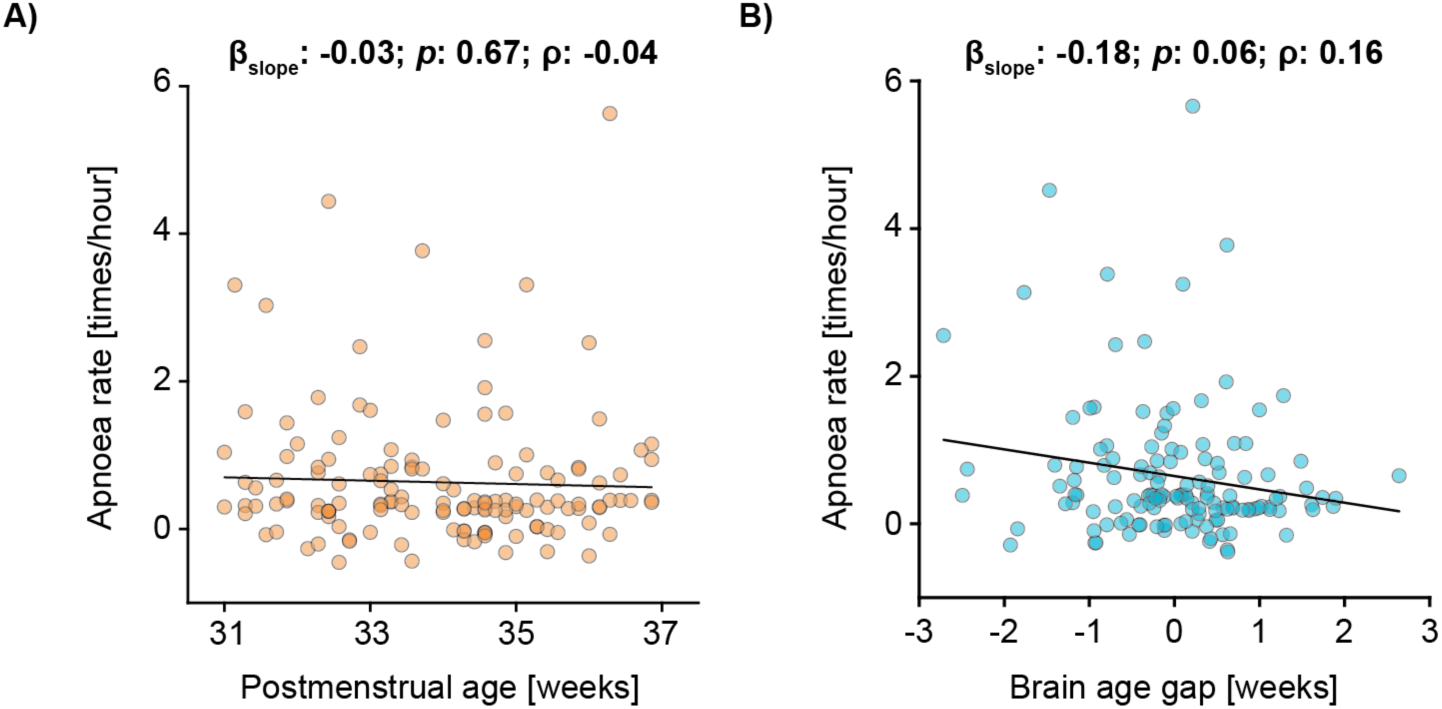
Apnoea rate against postmenstrual age and brain age gap with the linear regression models adjusted for data length, suspected infection and mode of ventilation. Apnoea rate associations with **A)** postmenstrual age (PMA) and **B)** brain age gap. Brain age gap is computed relative to the age of the infant, with negative values indicating that the brain activity is immature relative to the infant’s PMA, and positive values indicating that the brain activity is more mature relative to the infant’s PMA. Each dot indicates an individual test occasion (74 infants were studied on 138 test occasions). Black line is the mean fit of the linear model. Regression models are adjusted for infection, data length of inter-breath intervals and mode of ventilation.

**Figure S12.**
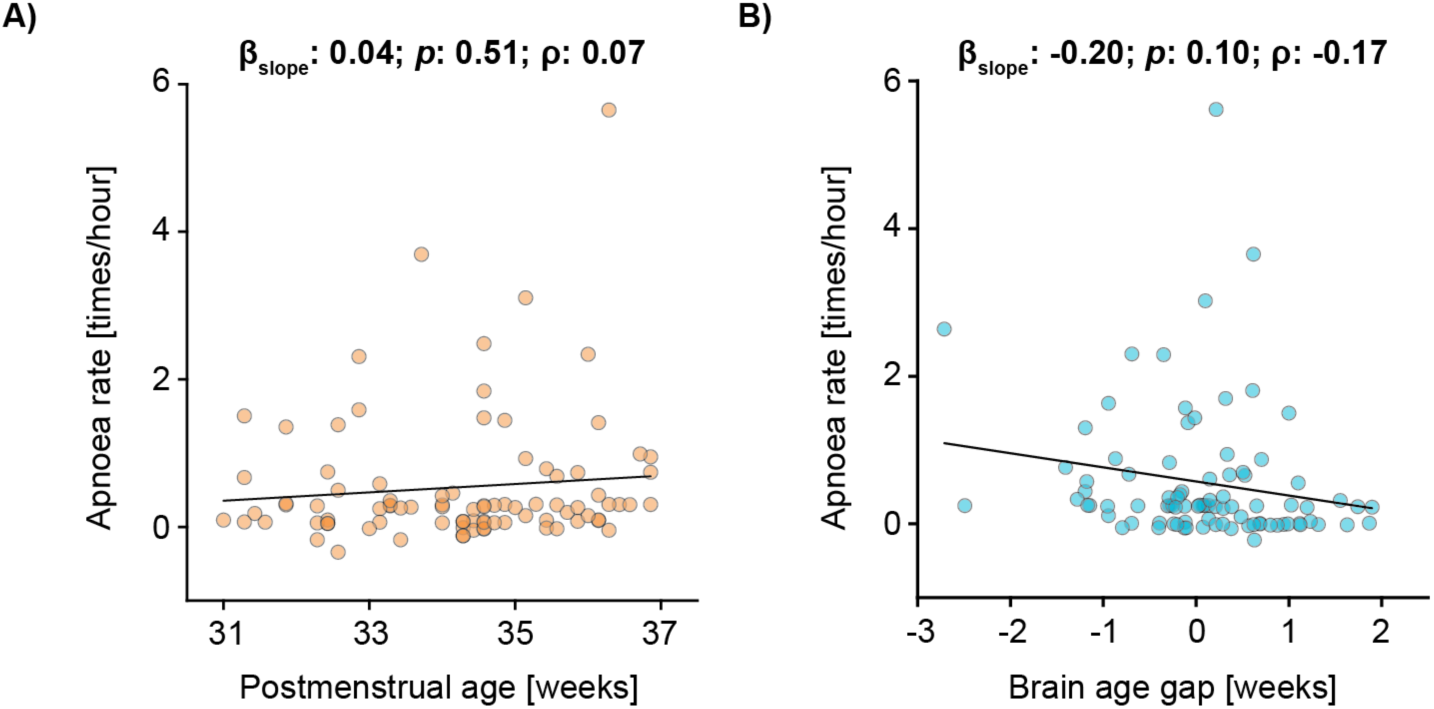
Apnoea rate against postmenstrual age and brain age gap for self-ventilating babies. Apnoea rate associations with **A)** postmenstrual age (PMA) and **B)** brain age gap. Brain age gap is computed relative to the age of the infant, with negative values indicating that the brain activity is immature relative to the infant’s PMA, and positive values indicating that the brain activity is more mature relative to the infant’s PMA. Each dot indicates an individual test occasion (57 infants were studied on 89 test occasions). Black line is the mean fit of the linear model. Regression models are adjusted for infection and data length of inter-breath intervals.

**Figure S13.**
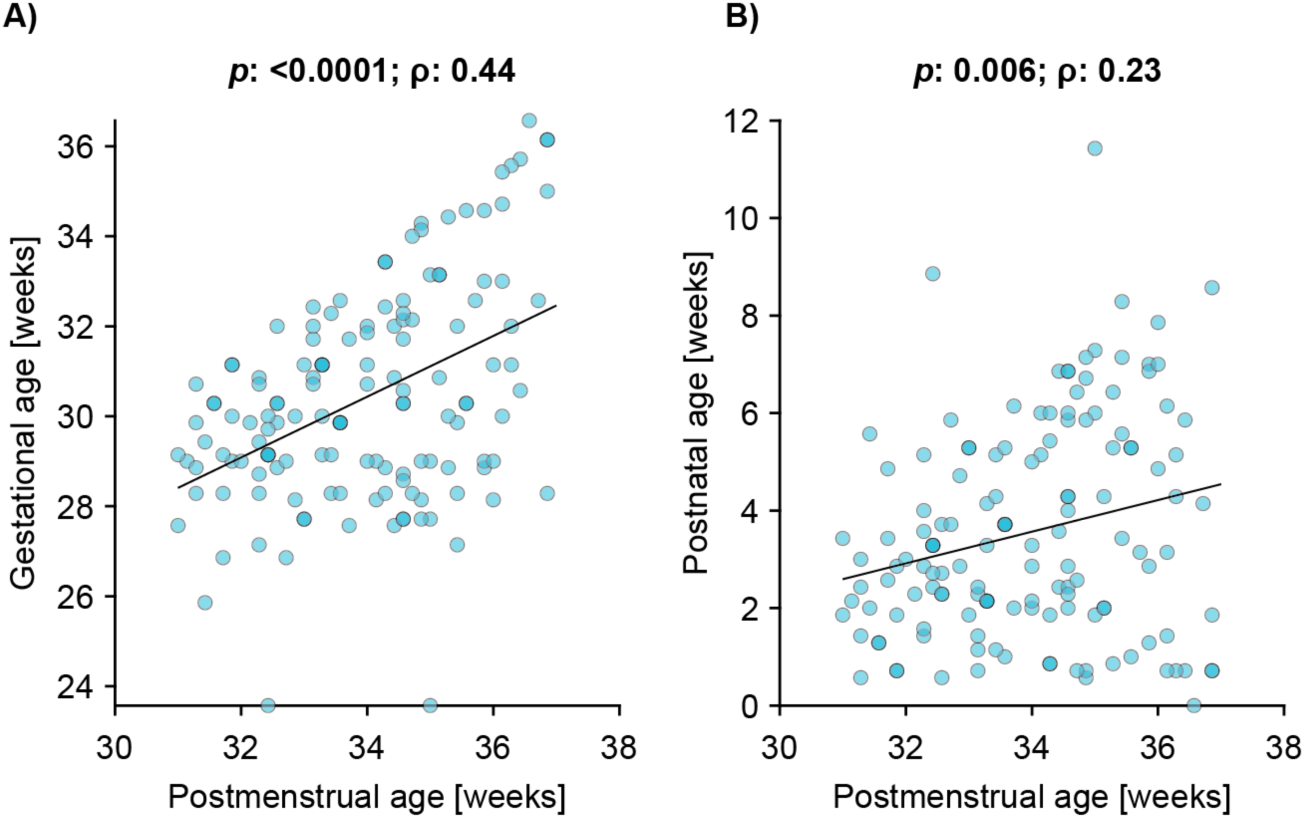
Correlations between A) postmenstrual age and gestational age, and B) postmenstrual age and postnatal age.

**Figure S14.**
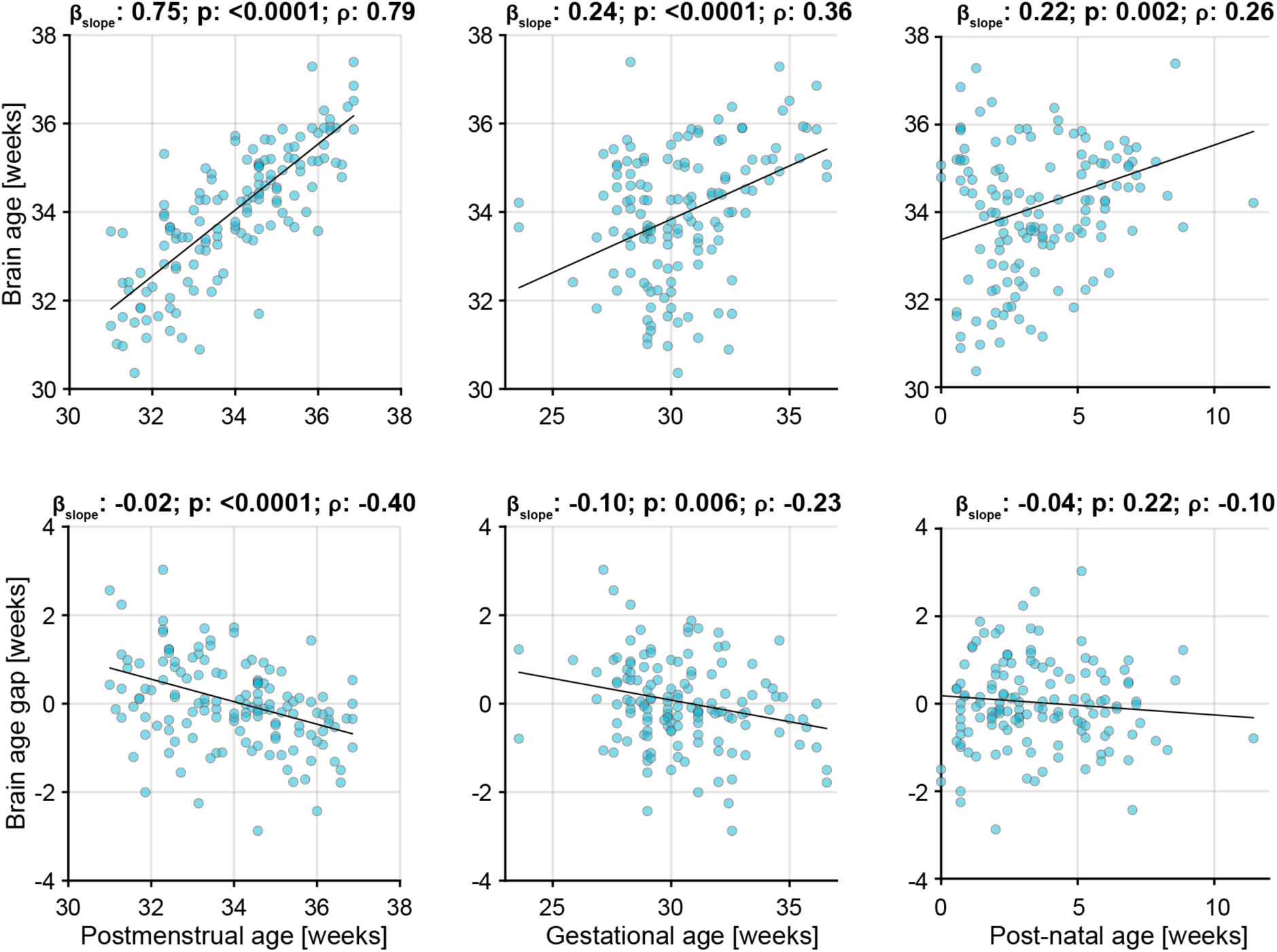
Associations between brain age (gap) with postmenstrual age, gestational age and post-natal age. Brain age is estimated from both the resting state and sensory model. The black graph is the mean fit of a linear mixed effects model. Brain age gap is computed relative to the age of the infant, with negative values indicating that the brain activity is immature relative to the infant’s PMA, and positive values indicating that the brain activity is more mature relative to the infant’s PMA. Each dot indicates an individual test occasion.

**Figure S15.**
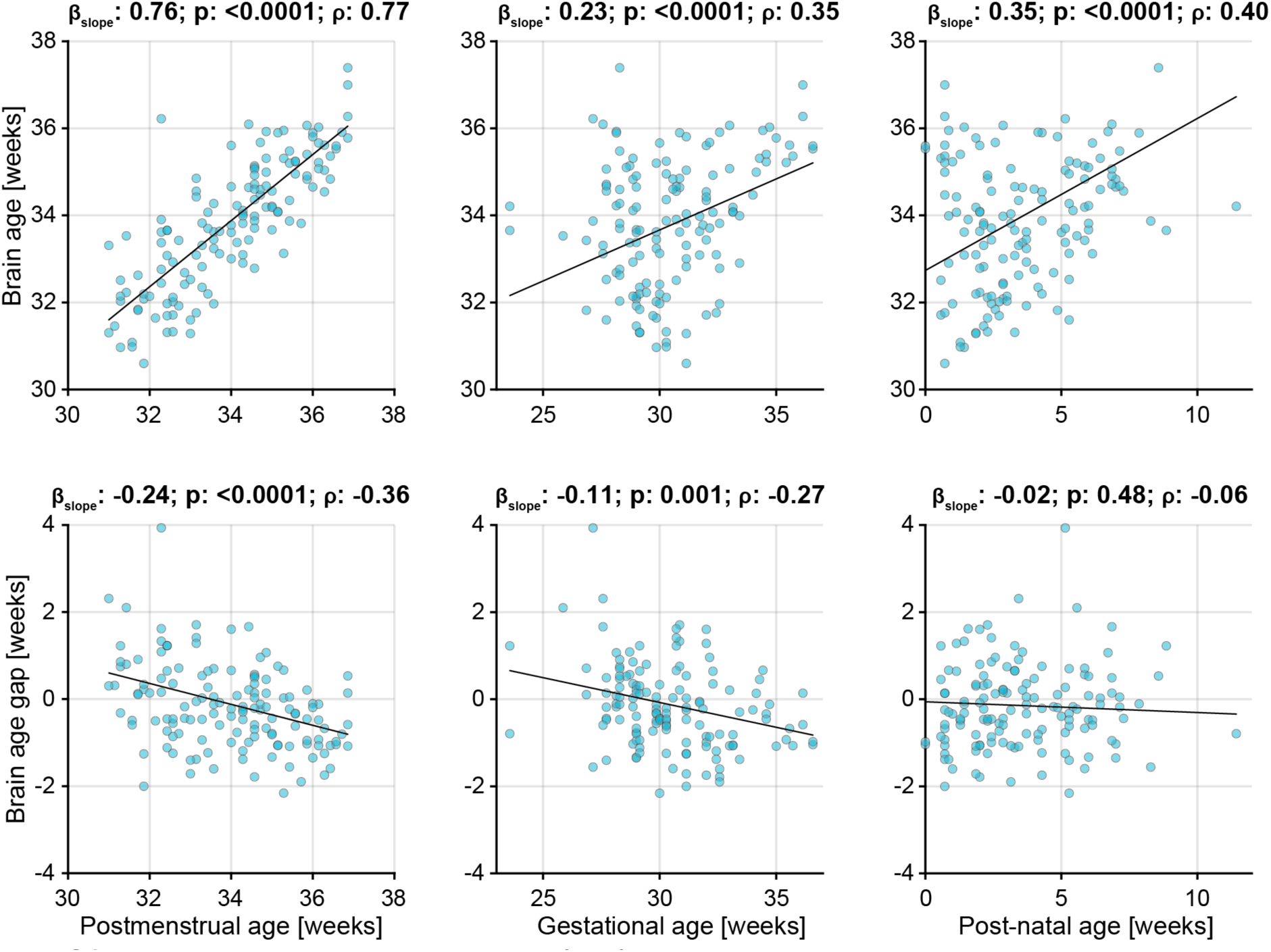
Associations between brain age (gap) with postmenstrual age, gestational age and post-natal age using the resting state model. Brain age is estimated from the resting state model. The black graph is the mean fit of a linear mixed effects model. Brain age gap is computed relative to the age of the infant, with negative values indicating that the brain activity is immature relative to the infant’s PMA, and positive values indicating that the brain activity is more mature relative to the infant’s PMA. Each dot indicates an individual test occasion.

**Figure S16.**
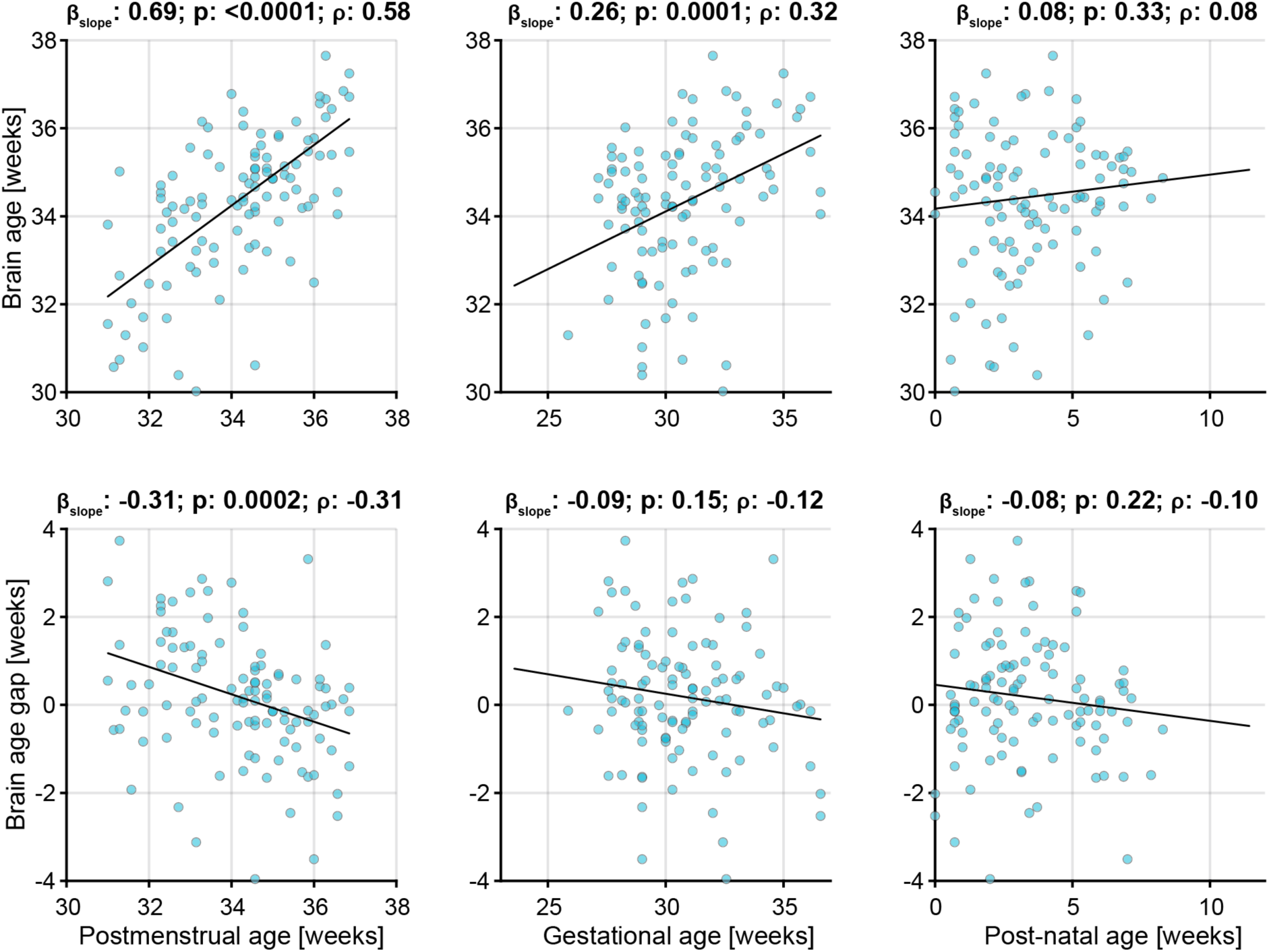
Associations between brain age (gap) with postmenstrual age, gestational age and post-natal age using the sensory-evoked activity. Brain age is estimated from the sensory model. The black graph is the mean fit of a linear mixed effects model. Brain age gap is computed relative to the age of the infant, with negative values indicating that the brain activity is immature relative to the infant’s PMA, and positive values indicating that the brain activity is more mature relative to the infant’s PMA. Each dot indicates an individual test occasion.

## Notes

### Competing Interest Statement

The authors have declared no competing interest.

### Funding Statement

This study was funded by the Wellcome Trust and Royal Society through a Sir Henry Dale Fellowship awarded to CH (grant number: 213486/Z/18/Z). FU is funded by the Commonwealth Scholarship Commission.

### Author Declarations

The UK National Research Ethics Service gave ethical approval for this work.

### Summary of Updates

Wording has been updated for clarity compared with other literature. In particular, the term 'brain maturity' has been revised to 'brain age gap' to maintain consistency with other published work in the field. Additional clarification on the possible confounds and limitations has also been added.

