## Supplementary Material for "Brain activity as a candidate biomarker for personalised caffeine treatment in premature neonates"

ventilating at the time of study. Apnoea rate and respiratory rate are also related to mode of ventilation and so mode of ventilation may act as a mediator variable (rather than a confound variable). We therefore chose not to include it in the main statistical analysis but have also assessed the results with inclusion of this factor (Figure S11). Apnoea rate reveals a strong trend with brain age gap, but not with PMA (brain age gap: *p*: 0.06; $\beta$ [95% CI]: -0.18 [-0.37; 0.01]; $\rho$: -0.16; PMA: *p*: 0.67; $\beta$ [95% CI]: -0.03 [-0.15; 0.10]; $\rho$: -0.04). Moreover, to further validate the main results, we conducted a subgroup analysis with self-ventilating infants (Figure S). This demonstrated a correlation in the same direction between apnoea rate and brain age gap (*p*: 0.10; $\beta$ [95% CI]: -0.20 [-0.44; 0.04]; $\rho$: -0.17; PMA: *p*: 0.51; $\beta$ [95% CI]: 0.04 [-0.09; 0.18]; $\rho$: -0.07) but due to the lower sample size in this group (n=57 infants on 89 occasions) we were likely underpowered to demonstrate a significant effect.

We considered gestational age (age in weeks at birth) and postnatal age (expressed as age in weeks after birth). Both are highly correlated with PMA (age in weeks at the time of study, which is the sum of gestational age and postnatal age) (Figure S13) and we, hence, did not add these to the model due to issues with collinearity.

Linearity of factors

We tested the assumption of linearity for the linear models (Figure 2A, B, D, E, and G) using multivariable fractional polynomial models. These models determine whether relationships are better modelled through non-linear relationships by finding the most appropriate functional form for every continuous predictor. Using the mfp package in R, we estimated these most appropriate forms and statistically tested them against the factor being included as linear. We included the same factors as presented in the main text (Figure 2A, B, D, E, and G; i.e., infection and data length when estimating apnoea rate and infection for respiration rate). For all models, infant was added as random factor. The p-values of Table S1 represents the difference in deviances between the fractional polynomial and linear models. As indicated by this table, none of the p-values for any model were statistically significant, confirming the linearity assumption.

Table S1. Statistical output of fractional polynomials relative to linear models

| **Main text figure** | **Predictor** | **Response** | **P-value** |
|---|---|---|---|
| 2A | Postmenstrual age | Apnoea rate | 0.49 |
| 2B | Brain age gap | Apnoea rate | 0.67 |
| 2D | Postmenstrual age | Respiration rate | 0.50 |
| 2E | Brain age gap | Respiration rate | 0.93 |
| 2G | Brain age gap | PMA at caffeine cessation | 0.99 |

*Exploratory hypothesis-generating findings*

In the main text we identified a possible relationship between brain age gap and apnoea rate in the week after caffeine treatment is discontinued, in an exploratory hypothesis-generating study. For a power calculation for future studies, based on the apnoea rates from our analyses, we obtained an effect size of 1.19 using these values in G*power 3.1, and together with an $\alpha$ of 0.05 and power of 0.95, the required sample size is $n$ = 34 for both groups (i.e., 17 participants per group).

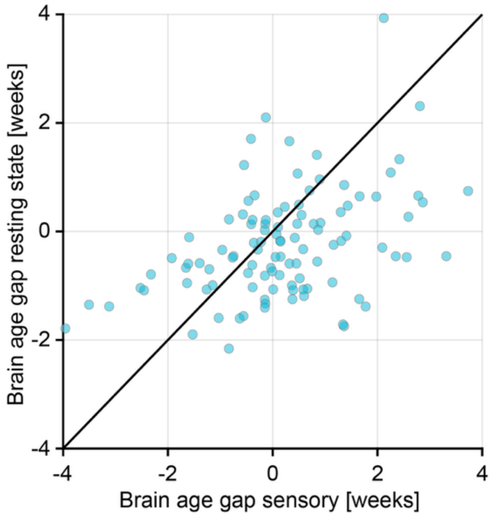


**Figure S1. Brain age gap estimates from the two brain age models (computed from the sensory-evoked and resting state EEG activity).** Brain age gap is computed relative to the age of the infant, with negative values indicating that the brain activity is immature relative to the infant's PMA, and positive values indicating that the brain activity is more mature relative to the infant's PMA. Each dot indicates an individual test occasion.

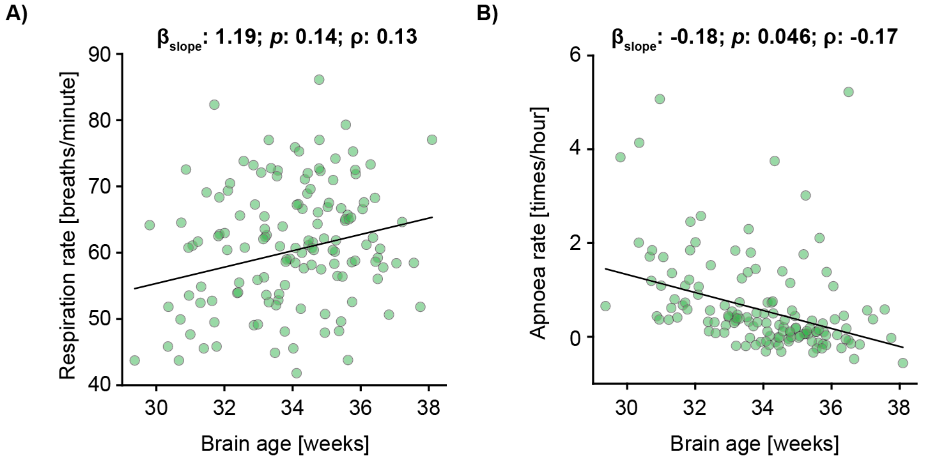


**Figure S2. Respiration and apnoea rate compared with brain age.** Brain age associations with **A)** respiration rate and **B)** apnoea rate. Brain age gap (as presented in the rest of the text) is defined as the difference in brain age minus postmenstrual age (PMA) (i.e. brain age is PMA plus brain age gap). Each dot indicates an individual test occasion (74 infants were studied on 138 test occasions). Black line is the mean fit of the linear model. Regression models are adjusted for infection, data length of inter-breath intervals, and postmenstrual age.

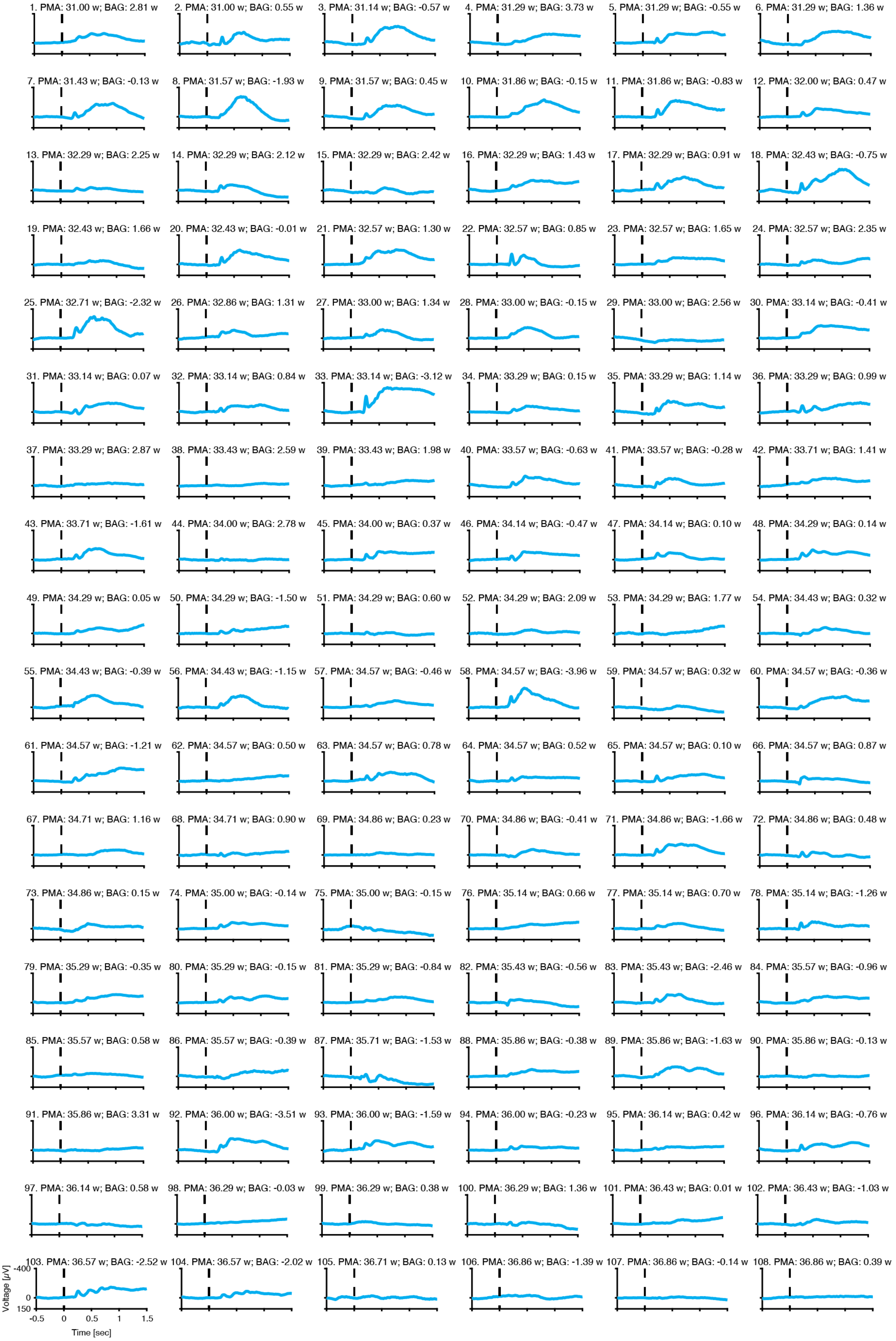


**Figure S3. Recording-specific evoked potentials for the visual stimuli.** Average (across all visual stimuli) evoked response for each individual recording, with the infants age and brain age gap indicated in the title of each subplot. Recordings are ordered according to increasing postmenstrual age (PMA). Stimulus was applied at Time = 0 sec. BAG: brain age gap; w: weeks.

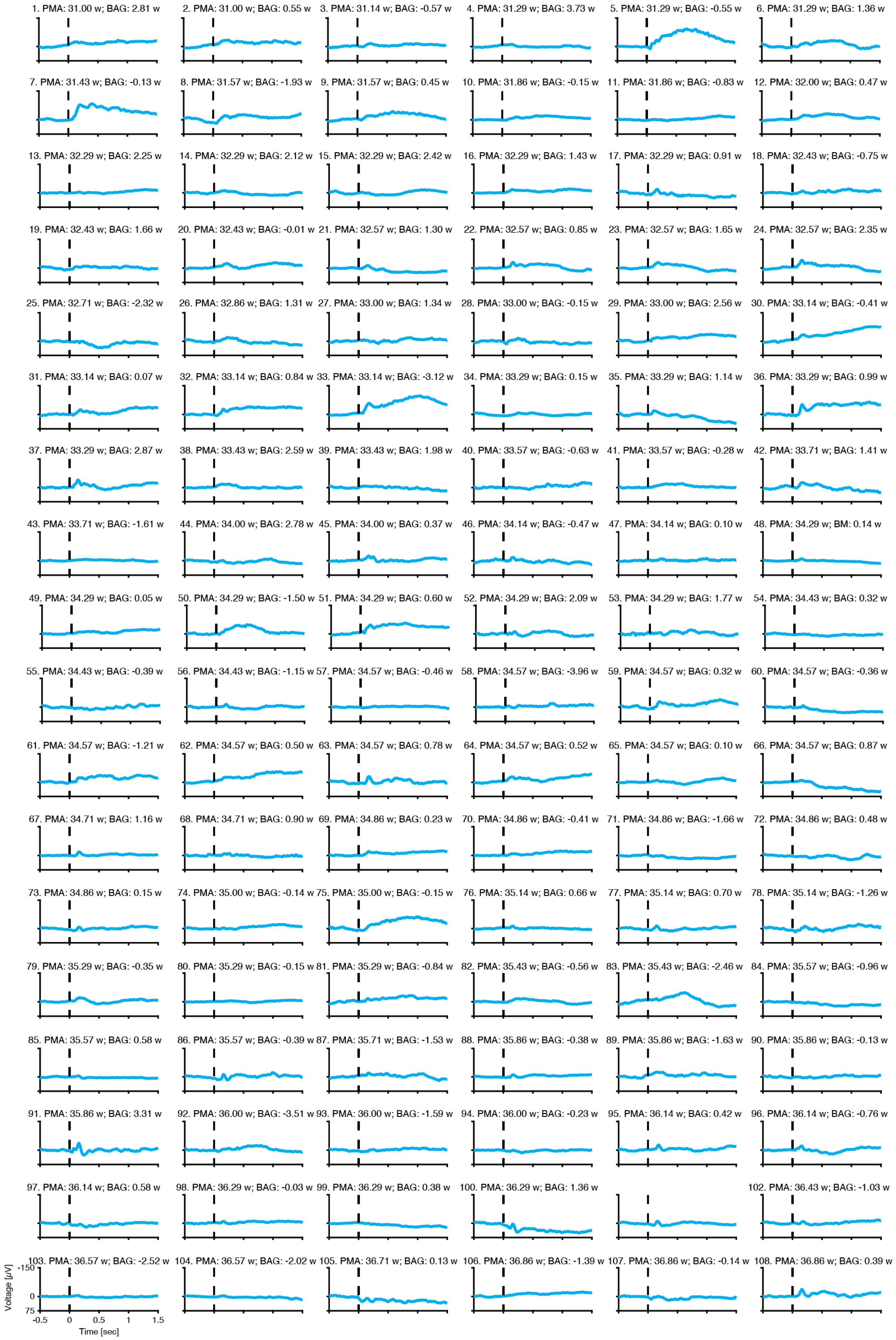


**Figure S4. Recording-specific evoked potentials for the tactile stimuli.** Average (across all visual stimuli) evoked response for each individual recording, with the infants age and brain age gap indicated in the title of each subplot. Recordings are ordered according to increasing postmenstrual age (PMA). Stimulus was applied at Time = 0 sec. BAG: brain age gap; w: weeks.

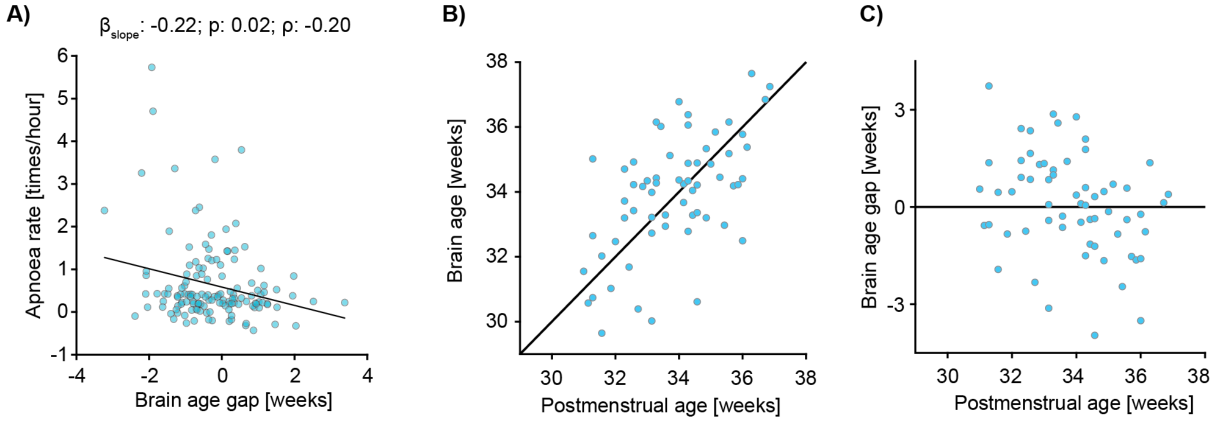


**Figure S5. Validation of the sensory-evoked brain age model.** The sensory-evoked model developed by Zandvoort et al. had an overlap with the data presented in this paper of 59 [out of 128] recordings from 48 babies. To check that this did not affect the results presented in the main text we investigated **A)** the relationship between brain age gap and apnoea rate excluding the sensory brain age estimates of babies that were part of the training set in Zandvoort et al. This showed similar results. Further, we validated the sensory-evoked model on the data that was not part of the original training set. **B-C**) Sensory brain age (gap) relationships with postmenstrual age for the data that was not included in the training set. The mean absolute error for this data was 1.22 weeks which is comparable to the mean absolute error of 1.41 weeks for the training dataset in the original paper.

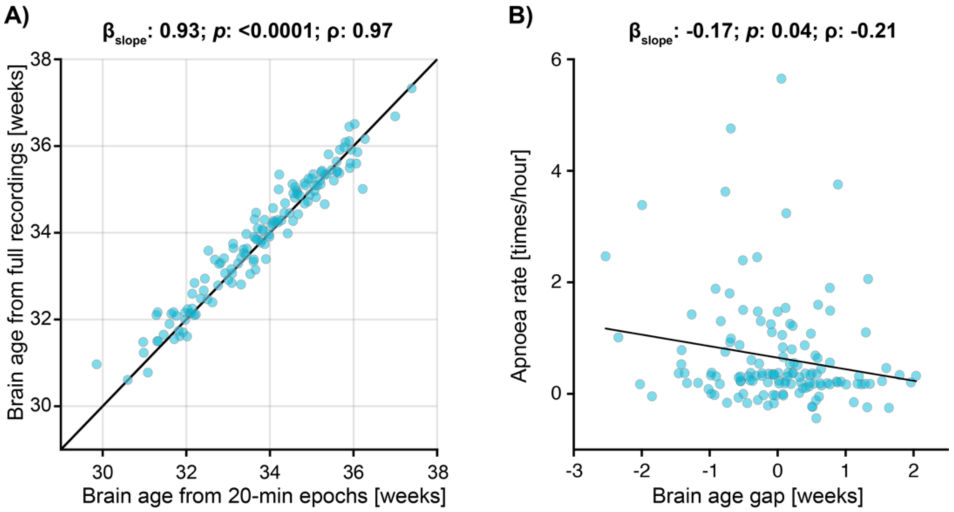


**Figure S6. Effects of data length on resting state brain ages. A)** Resting state brain ages predicted by 20-minutes epochs against those estimated over full recordings. The 20-minute epochs were used in the main results as this provided a consistent data length across all recordings and was the minimum data length which can be reliably used by the model as identified by Ansari et al.[14] **B)** Apnoea rate association with brain age gap when full recordings are used to estimate resting state brain age. The black line is the mean fit of a linear mixed effects model. Brain age gap is computed relative to the age of the infant, with negative values indicating that the brain activity is immature relative to the infant's PMA, and positive values indicating that the brain activity is more mature relative to the infant's PMA. For both panels, each dot indicates an individual test occasion.

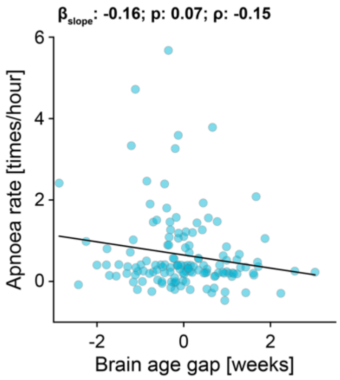


**Figure S7. Effects of bias correction on resulting association between apnoea rate and brain age gap.** Brain age gap is estimated from both resting state and sensory model. However, here we did not correct for deviations between brain age and PMA, which allowed for correlations between brain age gap and PMA. This still resulted in a significant relationship between brain age gap and apnoea rate. The black graph is the mean fit of a linear mixed effects model. Brain age gap is computed relative to the age of the infant, with negative values indicating that the brain activity is immature relative to the infant's PMA, and positive values indicating that the brain activity is more mature relative to the infant's PMA. Each dot indicates an individual test occasion.

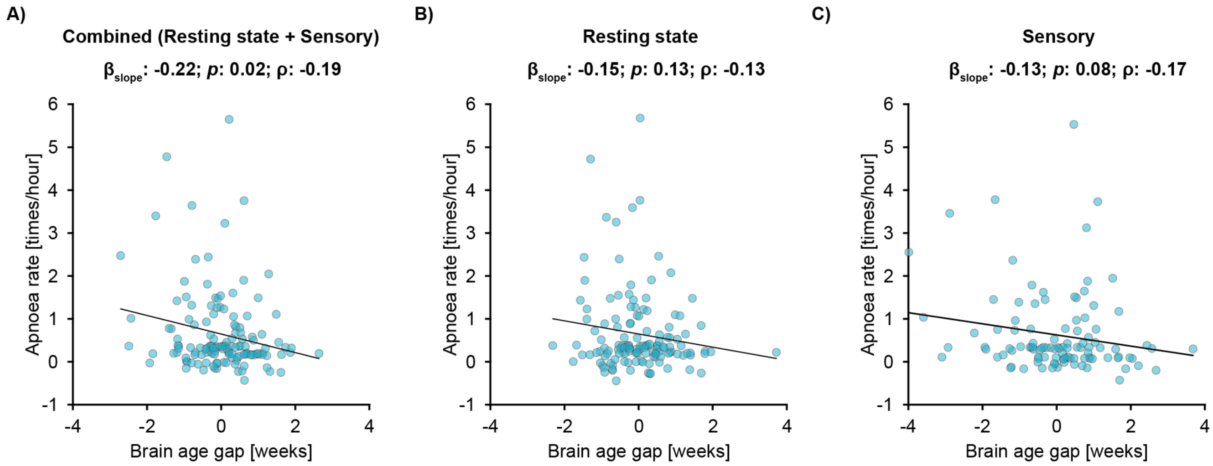


**Figure S8. Apnoea rate against brain age gap using the brain ages from different models.** Apnoea rate associations with **A)** both models, **B)** resting state model, and **C)** sensory model. Brain age gap is computed relative to the age of the infant, with negative values indicating that the brain activity is immature relative to the infant's PMA, and positive values indicating that the brain activity is more mature relative to the infant's PMA. Each dot indicates an individual test occasion. Black line is the mean fit of the linear model. Regression models are adjusted for infection and data length of inter-breath intervals.

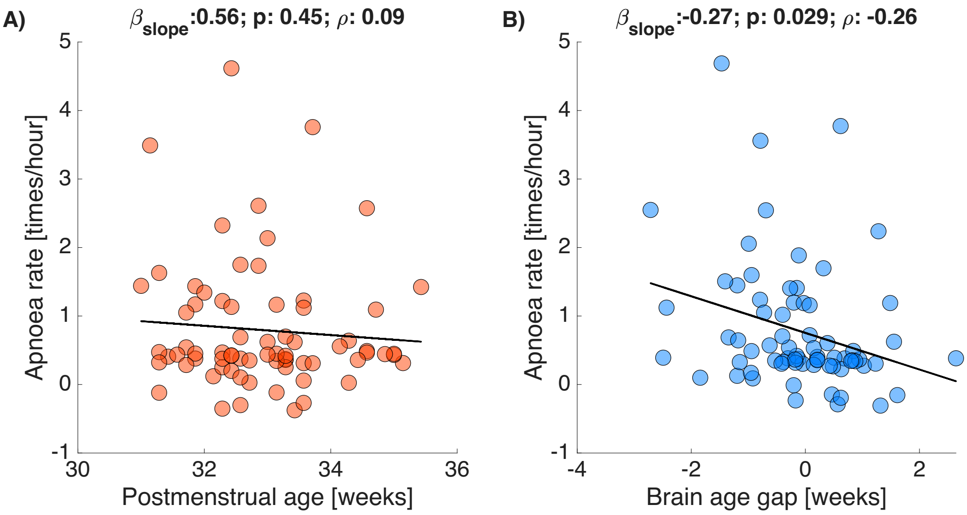


**Figure S9. Apnoea rate against postmenstrual age and brain age gap for infants receiving caffeine at the time of study.** Apnoea rate associations with **A)** postmenstrual age (PMA) and **B)** brain age gap. Brain age gap is computed relative to the age of the infant, with negative values indicating that the brain activity is immature relative to the infant's PMA, and positive values indicating that the brain activity is more mature relative to the infant's PMA. Each dot indicates an individual test occasion (40 infants were studied on 71 test occasions). Black line is the mean fit of the linear model. Regression models are adjusted for infection and data length of inter-breath intervals.

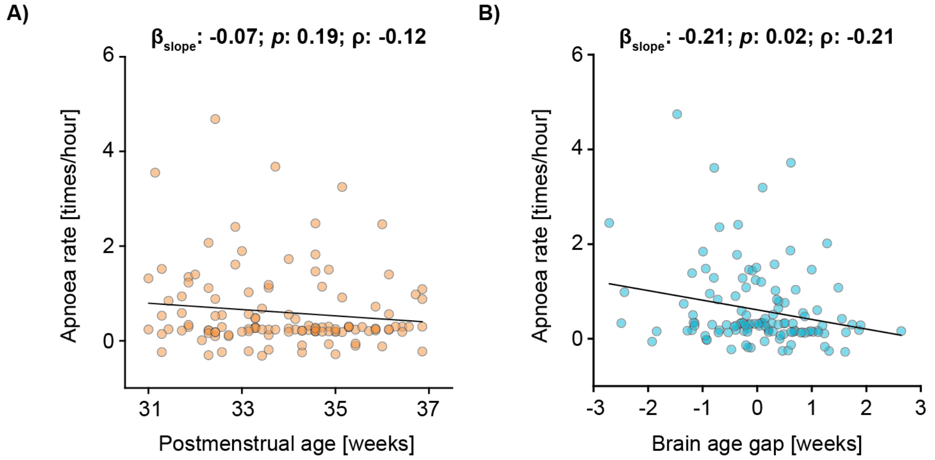


**Figure S10. Apnoea rate against postmenstrual age and brain age gap for infants without infection at the time of study.** Apnoea rate associations with **A)** postmenstrual age (PMA) and **B)** brain age gap. Brain age gap is computed relative to the age of the infant, with negative values indicating that the brain activity is immature relative to the infant's PMA, and positive values indicating that the brain activity is more mature relative to the infant's PMA. Each dot indicates an individual test occasion (69 infants were studied on 121 test occasions). Black line is the mean fit of the linear model. Regression models are adjusted for infection and data length of inter-breath intervals.

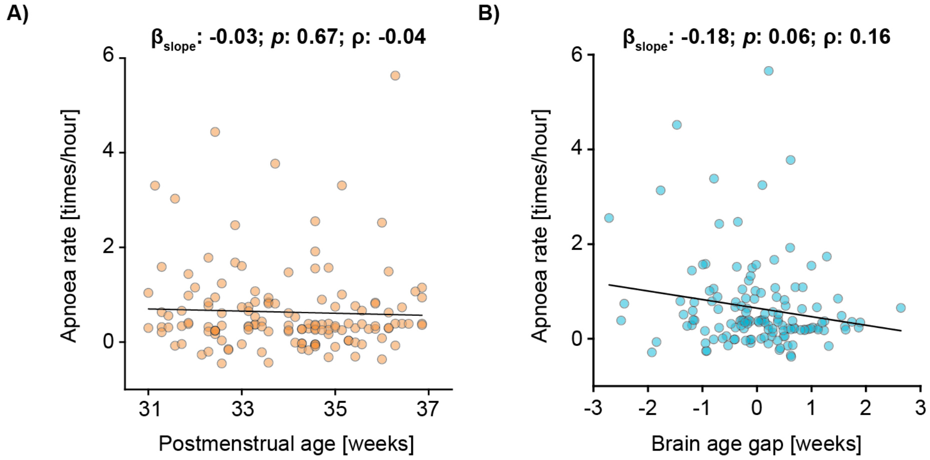


**Figure S11. Apnoea rate against postmenstrual age and brain age gap with the linear regression models adjusted for data length, suspected infection and mode of ventilation.** Apnoea rate associations with **A)** postmenstrual age (PMA) and **B)** brain age gap. Brain age gap is computed relative to the age of the infant, with negative values indicating that the brain activity is immature relative to the infant's PMA, and positive values indicating that the brain activity is more mature relative to the infant's PMA. Each dot indicates an individual test occasion (74 infants were studied on 138 test occasions). Black line is the mean fit of the linear model. Regression models are adjusted for infection, data length of inter-breath intervals and mode of ventilation.

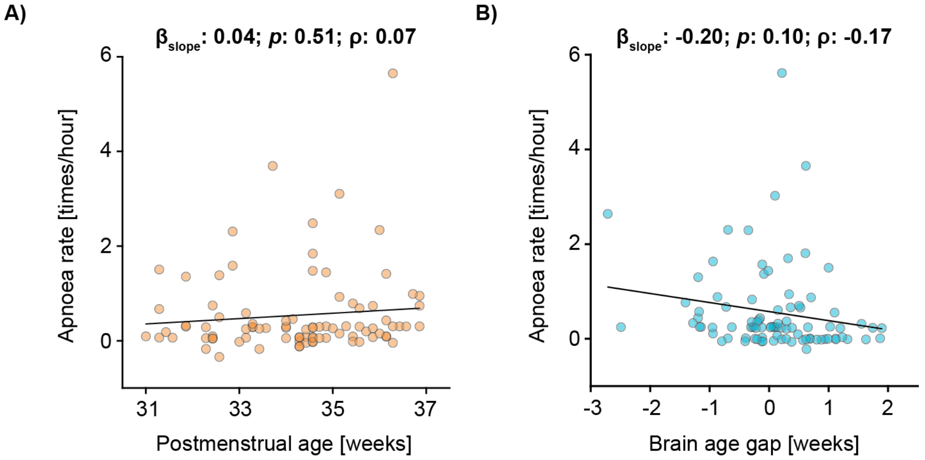


**Figure S12. Apnoea rate against postmenstrual age and brain age gap for self-ventilating babies.** Apnoea rate associations with **A)** postmenstrual age (PMA) and **B)** brain age gap. Brain age gap is computed relative to the age of the infant, with negative values indicating that the brain activity is immature relative to the infant's PMA, and positive values indicating that the brain activity is more mature relative to the infant's PMA. Each dot indicates an individual test occasion (57 infants were studied on 89 test occasions). Black line is the mean fit of the linear model. Regression models are adjusted for infection and data length of inter-breath intervals.

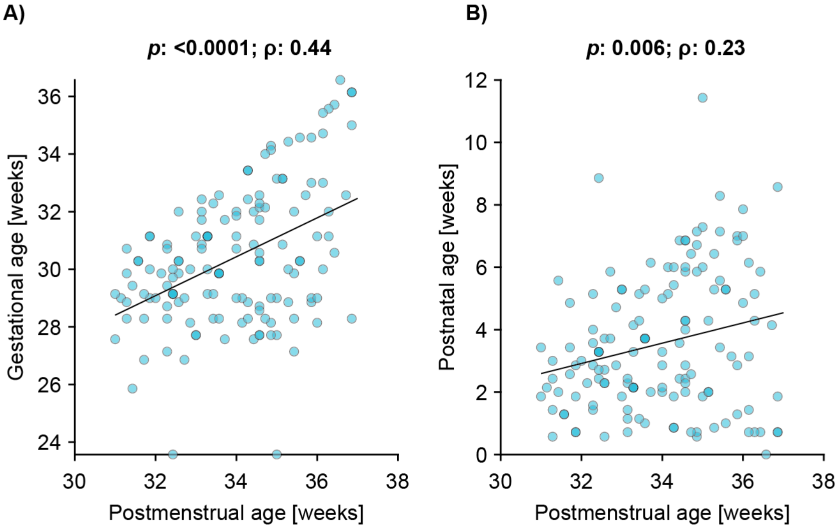


**Figure S13. Correlations between A) postmenstrual age and gestational age, and B) postmenstrual age and postnatal age.**

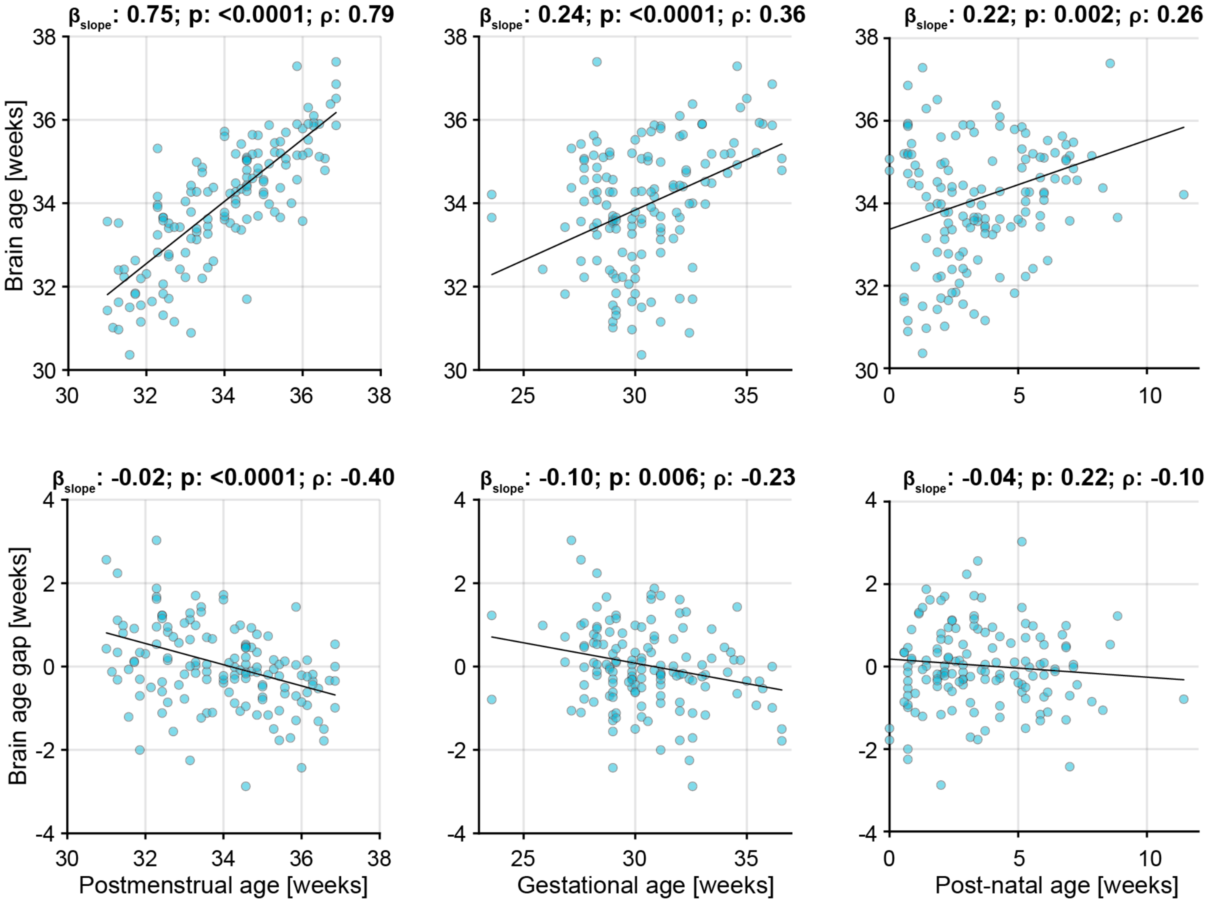


**Figure S14. Associations between brain age (gap) with postmenstrual age, gestational age and post-natal age.** Brain age is estimated from both the resting state and sensory model. The black graph is the mean fit of a linear mixed effects model. Brain age gap is computed relative to the age of the infant, with negative values indicating that the brain activity is immature relative to the infant's PMA, and positive values indicating that the brain activity is more mature relative to the infant's PMA. Each dot indicates an individual test occasion.

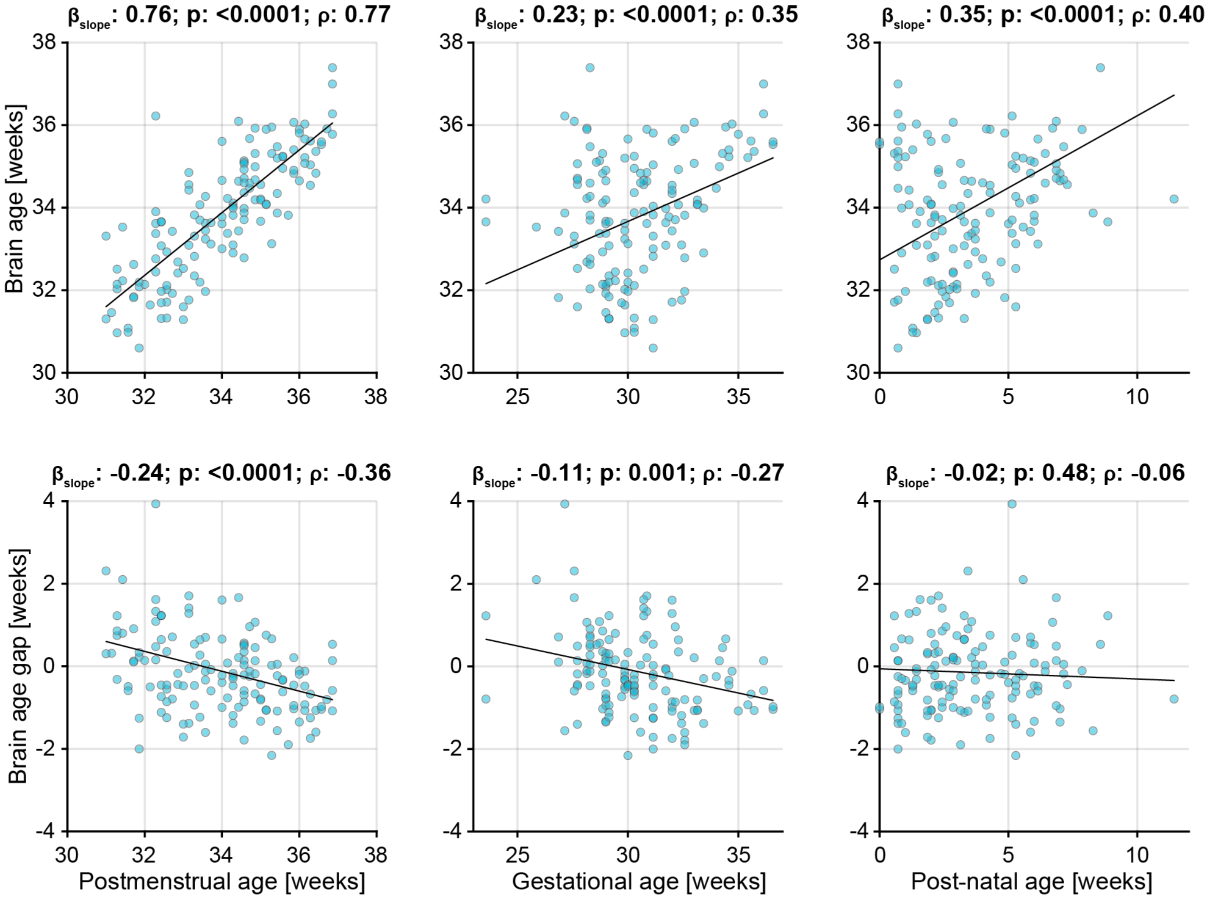


**Figure S15. Associations between brain age (gap) with postmenstrual age, gestational age and post-natal age using the resting state model.** Brain age is estimated from the resting state model. The black graph is the mean fit of a linear mixed effects model. Brain age gap is computed relative to the age of the infant, with negative values indicating that the brain activity is immature relative to the infant's PMA, and positive values indicating that the brain activity is more mature relative to the infant's PMA. Each dot indicates an individual test occasion.

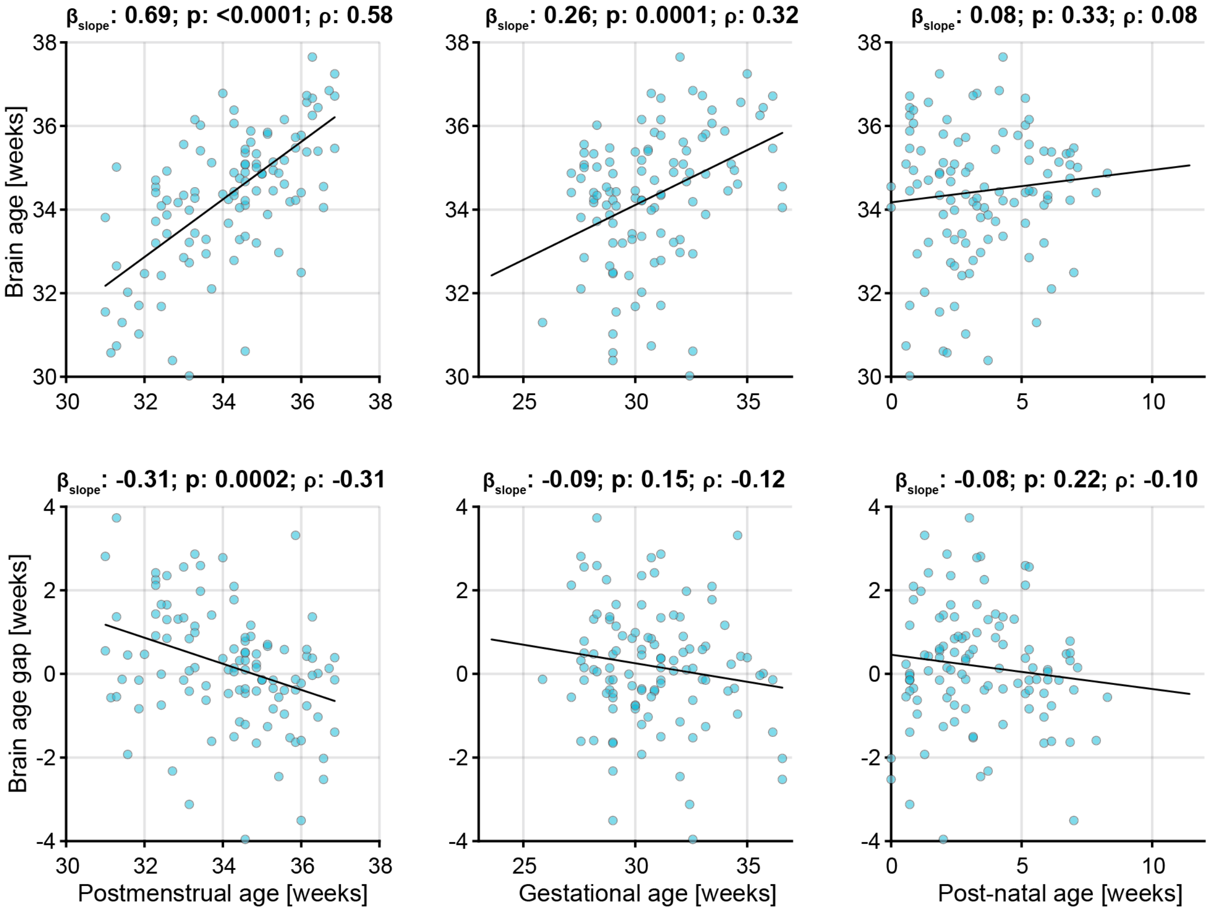


**Figure S16. Associations between brain age (gap) with postmenstrual age, gestational age and post-natal age using the sensory-evoked activity.** Brain age is estimated from the sensory model. The black graph is the mean fit of a linear mixed effects model. Brain age gap is computed relative to the age of the infant, with negative values indicating that the brain activity is immature relative to the infant's PMA, and positive values indicating that the brain activity is more mature relative to the infant's PMA. Each dot indicates an individual test occasion.
